# *Dsp^S311A^* knock-in mice replicate the clinical-pathological features of dominant and recessive forms of Desmoplakin-related cardiomyopathies

**DOI:** 10.1101/2025.01.14.24319713

**Authors:** Anna Di Bona, Anna Guazzo, Induja Perumal Vanaja, Riccardo Bariani, Maria C. Disalvo, Mattia Albiero, Nicolas Kuperwasser, Pierre David, Rudy Celeghin, Vittoria Di Mauro, Arianna Scalco, María López-Moreno, Monica De Gaspari, Mila Della Barbera, Stefania Rizzo, Domenico Corrado, Barbara Bauce, Giuseppe Zanotti, Gaetano Thiene, Kalliopi Pilichou, José Maria Perez Pomares, Mario Pende, Cristina Basso, Marco Mongillo, Tania Zaglia

**Author notes:** **Correspondence to:** Tania Zaglia, PhD, Department of Cardiac, Thoracic, Vascular Sciences and Public Health, University of Padova, via Giustiniani 2, 35128 Padova, Italy Department of Biomedical Science, University of Padova, via Ugo Bassi 58/B, 35131 Padova, Italy Veneto Institute of Molecular Medicine (VIMM), via Orus 2, 35129 Padova, Italy. equal contribution (Anna Di Bona, Anna Guazzo, Induja Perumal Vanaja).

## Abstract

**Background/Purpose:** Desmoplakin (DSP) mutations are linked to familial cardiomyopathies with a very high arrhythmogenic propensity. While autosomal recessive inheritance forms manifest in the cardio-cutaneous Carvajal syndrome, the dominant-inheritance variants associate to DSP-cardiomyopathy (DSP-CM). This latter is a subtype of Arrhythmogenic Cardiomyopathy characterized by frequent myocarditis-like episodes, dominant left ventricular (LV) remodeling, recurrent premature ventricular contractions and life-threatening arrhythmias, frequently preceding LV dysfunction and dilation. Notably, DSP-CM evades the diagnostic identifiers of Arrhythmogenic Cardiomyopathy, further complicating risk-stratification and prediction. At the time being, the pathogenetic mechanisms underlying DSP-related cardiomyopathies are largely obscure and their elucidation is urgently required.

**Methods:** To this end, we employed CRISPR-Cas9 to generate a novel knock-in mouse model harboring a point mutation at the murine ortholog of human Serine-299, a mutation site previously identified in a family affected by left dominant-Arrhythmogenic Cardiomyopathy. In both heterozygotes and homozygotes, cardiac function was assessed by echocardiography and telemetry-ECG, at different ages. Results were correlated with heart structure, which was assessed by ultrastructural, histopathological and molecular/biochemical assays. The effects of moderate exercise on disease manifestations were tested.

**Results:** The homo- and hetero-zygous expression of mutant *Dsp^S311A^* allele replicated the human cardiac phenotypes of Carvajal syndrome and DSP-CM, respectively. Indeed, *Dsp^S311A/S311A^* mice featured precocious dilated cardiomyopathy with biventricular fibrotic remodeling, aneurisms, systolic dysfunction, increased arrhythmic vulnerability, sudden death and, remarkably, cutaneous defects. Differently, *Dsp^WT/S311A^* mice did not show evident cutaneous alterations, and myocardial remodeling and contractile dysfunction developed later and were associated to increased cell death, inflammatory response and patchy fibrosis predominantly in the LV. Notably, as observed in certain patient subgroups, *Dsp^WT/S311A^* mice had electrophysiological alterations (i.e. QRS prolongation, distal conduction defects and sustained ventricular arrhythmias) prior to developing contractile dysfunction. Furthermore, in both genotypes, exercise accelerated myocardial remodeling and increased the incidence of arrhythmic mortality.

**Conclusions:** Our novel *Dsp^S311A^* mice recapitulate the clinical and pathological features of the respective dominant (i.e. DSP-CM) and recessive (i.e. Carvajal syndrome) forms of DSP-related cardiomyopathies. Thus, *Dsp^S311A^* mice are a novel experimental model of human diseases, suited to test therapeutic interventions aimed at reducing the burden of stress-dependent SD.

## INTRODUCTION

Desmoplakin Cardiomyopathy (DSP-CM) is a subtype of Arrhythmogenic Cardiomyopathy (AC), resulting from expression of pathogenic variants in the *DSP* gene. Such inherited cardiomyopathy is hallmarked by predominant involvement of the left ventricle (LV) with inflammation, fibrosis, increased susceptibility to stress-dependent ventricular arrhythmias and variable degree of contractile dysfunction ^1–5^. DSP is the most extensively characterized plakin protein and plays critical roles in several biological processes, spanning from regulation of cardiac morphogenesis to development of both nervous and vascular systems^6^. Alongside Plakophilin-2 (PKP2), Plakoglobin (JUP), Desmocollin-2 (DSC2) and Desmoglein-2 (DSG2), DSP is a component of desmosomes, which connect adjacent cardiomyocytes, withstanding the mechanical stress of contraction ^1–5,7^. The importance of DSP in organismal homeostasis is underscored by the identification of both autosomal dominant and recessive variants as primary causes of a range of cutaneous disorders, which may manifest in combination of- or without-cardiac abnormalities ^1,2,7–10^. Among the cardiac manifestations linked to *DSP* variants, Carvajal syndrome is an inherited disorder, caused by homozygous expression of two mutant *DSP* alleles, hallmarked by dilated cardiomyopathy, with diffuse scarring of both the right-(RV) and left (LV) ventricles and aneurysms, associated with palmoplantar keratoderma and woolly hair ^7–10^. Since its initial identification, numerous *DSP* variants associated with pathological cardiac remodeling, highly arrhythmic propensity and sudden death (SD) have been documented ^1–5^. Homo- or hetero-zygous expression of *DSP* variants share with AC the common denominator of non-ischemic injury of the ventricular myocardium with fibrous or fibro-fatty repair ^7–13^. Although the clinical spectrum displays considerable variability, notable features include: LV remodeling—either in isolation or accompanied by RV involvement; recurrent premature ventricular contractions (PVCs); frequent myocarditis-like presentation with chest pain and elevated Troponin-I levels in the absence of coronary disease (referred to as "hot-phase" episodes) ^2–5,14,15^. Importantly, clinical studies indicate that arrhythmias in DSP-CM often manifest in the absence of heart failure or significant LV systolic dysfunction ^1–5,14,15^. Notably, the 2010 diagnostic criteria for AC and the variables commonly employed for risk estimation (in both Dilated Cardiomyopathy and AC) fall short of sensitivity for predicting arrhythmic risk in DSP-CM ^16^. Thus, elucidating the mechanistic underpinnings of DSP-CM is critical to refine diagnostic strategies and reduce the burden of SD. To this end, the development of preclinical models that closely resemble human genetics and clinical manifestations is urgently required. Given that DSP is expressed in a wider array of cell types beyond cardiomyocytes ^6^, the generation of knock-in mice offers an appropriate model for comprehensive investigation of disease mechanisms in DSP-CM. In this study, we thus employed CRISPR-Cas9 to introduce a point mutation at the murine ortholog of human Serine-299, a mutation site previously identified in a family affected by LV-dominant AC ^1,2^. Phenotyping of cardiac function and structure of both homozygous and heterozygous *Dsp^S311A^* mice reflected faithfully the respective clinical-pathological manifestations of human Carvajal syndrome and DSP-CM. Furthermore, our findings show the effects of exercise on arrhythmic mortality and myocardial remodeling. The established characteristics of the *Dsp^S311A^* knock-in mouse make it a valuable disease model that adds insight into the relevant pathophysiological mechanisms underlying inherited arrhythmic syndromes and provides a robust platform for experimental investigations and drug development research.

## METHODS

### Human samples

We analysed a heart from a young male individual (age range 10-15 years) carrying the *Dsp^S299R^* variant and suddenly died at rest. The heart was acquired during routine post-mortem clinical investigations and archived at the Cardiovascular Pathology Unit of the University Hospital of Padova. The sample was used in accordance with the ‘Recommendation (CM/Rec (2016) 6) of the Committee of Ministers of the Member States on research on biological materials of human origin’, released by the European Council as received by the Italian National Council of Bioethics which reviewed and confirmed adherence to the principles outlined in the recommendation, ensuring that the dignity, privacy, and rights of individuals were fully protected. Heart samples were analyzed using protocols as previously described ^17,18^.

### Animal models

We here used newly generated knock-in mice carrying a point variant in *Dsp* (*Dsp^WT/S311A^* and *Dsp^S311A/S311A^*) and littermate controls (*Dsp^WT/WT^*). Both male and female mice were used at different ages (from 1 to 16 months). Animals were maintained in an Authorized Animal Facility (authorization number 175/2002A), at controlled temperature, with a 12-on/12-off light cycle and had access to water and food *ad libitum*. All experimental procedures were approved by the University of Padova ethical committee and by the Italian Ministry of Health (Authorization numbers 408/2018PR and 129/2018PR), in compliance with Italian Animal Welfare Law (Law n.116/1992 and subsequent modifications). Experiments were performed by trained personnel with documented formal training and previous experience in experimental animal handling and care. All procedures were refined prior to starting the study, and the number of animals was calculated to use the least number of animals sufficient, according to statistical sample power calculation.

### Generation of *Dsp^S311A^* knock-in mice

*Dsp^S311A^* knock-in mice were generated with the aid of the Transgenesis Platform, Laboratoire d’Expérimentation Animale et Transgenèse of Imagine Institute (Structure Fédérative de Recherche Necker). All animal procedures were performed with approval from the French Ministry of Research. Superovulation of four-weeks old C57BL6/J female mice was induced by intraperitoneal injections of 5 IU PMSG (SYNCRO-PART® PMSG 600 UI, Ceva), followed by 5 IU hCG (Chorulon 1500 UI, Intervet) at an interval of 36-48h. Female were mated with C57BL/6J male mice. Zygotes were collected from the oviducts of one-day fertilized mice, exposed to hyaluronidase (H3884, Sigma-Aldrich) to remove the cells of the cumulus oophorus, and then incubated in M2 medium (M7167, Sigma-Aldrich). The sgRNAs (IDT DNA) were hybridized with the Cas9 protein (IDT) and injected into the pronucleus of the C57BL/6J zygotes. Surviving zygotes were placed in KSOM medium (MR-106-D, Merck-Millipore) and cultured overnight to two-cell stage and transferred into the oviduct of B6CBAF1 pseudo-pregnant females (hybrid offspring of C57BL/6J females (B6) and CBA/J males (CBA)). The offspring DNA was validated by Sanger sequencing (Suppl.Fig.1D). All *Dsp*-mutant mice were backcrossed at least three times with C57BL/6J mice to remove potential off-targets. The Dsp-knock-in offspring were further confirmed by PCR genotyping with appropriate primers. By performing two injection sessions with Cas9 and Sg-RNA plus single stranded oligo donor nucleotide (ODN) for S311A, we obtained three *Dsp^WT/S311A^* founders out of 54 total mice born. Five of them were knock-out mice and the other were all wild-type mice. Heterozygous mice were viable and fertile and were thus backcrossed to expand the knock-in mouse line. Additional details are described in the **Supplementary Data** section.

**Figure 1.**
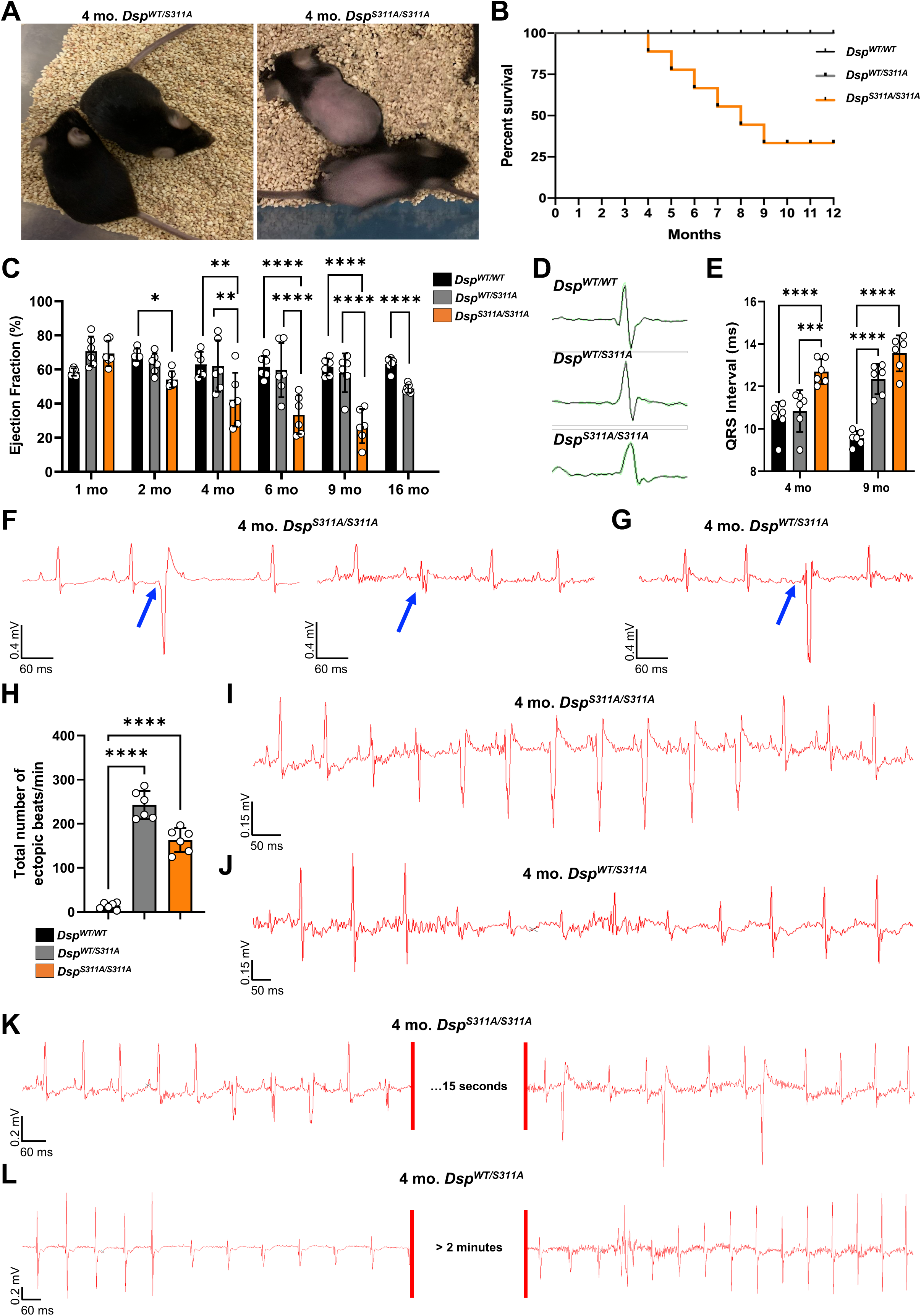
Cardiac functional phenotyping of *Dsp^S311A^* mice. (**A**) Image of 4 months old *Dsp^WT/S311A^* and *Dsp^S311A/S311A^* male mice. (**B**) Survival probability of male and female *Dsp^WT/WT^*, *Dsp^WT/S311A^* and *Dsp^S311A/S311A^* mice, estimated by Kaplan-Meier method. Comparison of survival curves was performed by both Mantel-Cox and Gehan-Breslow-Wilcoxon tests. (n>20 mice for each group). (**C**) Echocardiographic assessment of heart function in *Dsp^WT/WT^*, *Dsp^WT/S311A^* and *Dsp^S311A/S311A^* male mice, at different ages. Bars represent s.d. Differences among groups were determined using two-way ANOVA with Tukey’s test for multiple comparisons (for 1-to-9 months groups), and unpaired t-test for 16 months-old mice. (n=6 mice/group; *, p<0.05; **, p<0.01; ****, p<0.0001). (**D**) Representative ECG traces of QRS complex in 4 months *Dsp^WT/WT^*, *Dsp^WT/S311A,^* and *Dsp^S311A/S311A^* male mice. (**E**) QRS complex in *Dsp^WT/WT^*, *Dsp^WT/S311A^* and *Dsp^S311A/S311A^* male mice, averaged for 1000 beats/mouse. Bars represent s.d. Differences among groups were determined using two-way ANOVA with uncorrected Fisher’s LSD test for multiple comparisons. (n=6 mice/group; ***, p<0.001; ****, p<0.0001). (**F-G**) Representative ECG traces obtained in conscious homozygous (**F**) and heterozygous (**G**) mice. Blue arrows evidence ectopic beats. (**H**) ECG-based evaluation of the number of ectopic beats *per* minute, calculated in a ten minutes-continuous recording. Four months *Dsp^WT/WT^*, *Dsp^WT/S311A^* and *Dsp^S311A/S311A^* male mice were analyzed. Bars represent s.d. Differences among groups were determined using One-way ANOVA with Tukey’s test for multiple comparisons. (n=6 mice/group. ****, p<0.0001). (**I-L**) Representative ECG traces showing sustained arrhythmias in conscious homozygous (**I, K**) and heterozygous (**J, L**) mice.

### Statistics

Statistical analysis was performed using GraphPad Prism 8. Normality of data distribution was assessed with Shapiro-Wilk test. In datasets showing normal distribution, unpaired *t* test with Welch’s correction (comparison between two groups) or one-way analysis of variance (ANOVA) (comparison between three or more groups) were applied. For one-way ANOVA, depending on the result of SD equality (by a Brown–Forsythe test), ordinary or Brown–Forsythe and Welch ANOVA tests were performed. In cases where two factors were analyzed simultaneously (e.g. genotype and time points), a two-way ANOVA was used, followed by Tukey’s, Dunnett’s, Šídák’s or Uncorrected Fisher’s LSD test for multiple comparisons as appropriate. In non-normally distributed datasets, non-parametric tests were used. All data are expressed as the mean ± standard deviation (s.d.). Confidence intervals were set at 95% for statistical comparisons. A P value <0.05 was considered statistically significant.

Additional Methodological details are described in the **Supplementary Data** section.

## RESULTS

### Generation and phenotyping of *Dsp^S311A^* mice

We generated a murine model of DSP-CM by editing the *Dsp* gene to introduce a point mutation, resulting in the Alanine (A) substitution of amino acid Serine (S)-311 in the highly conserved DSP protein (*Dsp^S311A^*) (**Suppl.Fig.1A-C**). Ser311 is the murine ortholog of human S299, which was found mutated in a family diagnosed with a ‘left-dominant form of AC’ ^1,2^. *Dsp^S311A^* knock-in mice were generated by CRISPR/Cas9 which yielded three *Dsp^WT/S311A^* founders, backcrossed into the C57BL/6J background for at least 10 generations to expand the mouse line (see also **Suppl. Methods**). The offspring DNA was validated by Sanger sequencing (**Suppl.Fig.1D**), and genome sequencing excluded casual off target insertions (**Suppl. Methods** and **Suppl.Fig.2**). Both male and female *Dsp^WT/S311A^* and *Dsp^S311A/S311A^* knock-in mice were born at the expected Mendelian ratio, weaned normally and grew to the same body weight. At general examination, alopecia was evident in both male and female homozygous mice, from four months onwards (**Fig.1A**). Interestingly, while unstressed heterozygotes had normal lifespan, both male and female homozygous *Dsp* mutant mice showed decreased life expectancy due to elevated incidence of SD from 4 months onwards (**Fig.1B**).

**Figure 2.**
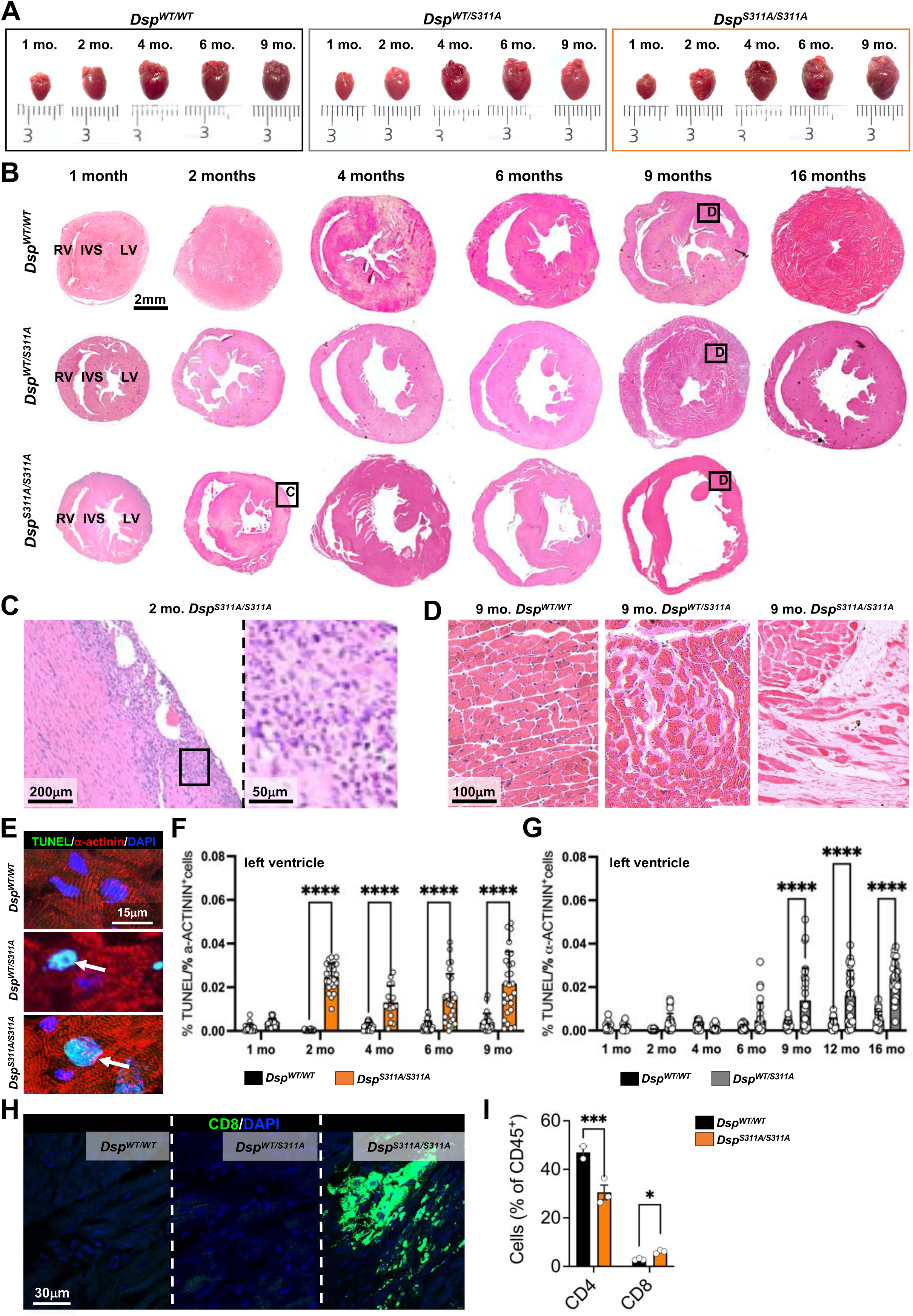
Cardiac structural phenotyping of *Dsp^S311A^* mice. (**A**) Hearts harvested from *Dsp^WT/WT^*, *Dsp^WT/S311A^* and *Dsp^S311A/S311A^* male mice at different ages. (**B**) Haematoxylin-eosin staining of sections from the mid portion of the ventricles of *Dsp^WT/WT^* (top panels), *Dsp^WT/S311A^* (middle panels) and *Dsp^S311A/S311A^* (bottom panels) male mice at different ages. RV, right ventricle; IVS, interventricular septum; LV, left ventricle. (**C**) Images are details of the LV subepicardial region of 2 months *Dsp^S311A/S311A^* heart (see black box in **B** bottom panel). (**D**) Images are details from the LV wall of 9 months *Dsp^WT/WT^*, *Dsp^WT/S311A^* and *Dsp^S311A/S311A^* hearts (see black boxes in **B**, top, middle and bottom panels). (**E**) TUNEL assay (green signal) in heart sections from 9 months *Dsp^WT/WT^, Dsp^WT/S311A^* and *Dsp*^S^*^311A/S311A^* male mice. Sections were co-stained with an antibody to α-actinin. Nuclei were counterstained with DAPI. Images are details from the LV. (**F-G**) Quantification of the number of TUNEL-positive cardiomyocytes in the LV from *Dsp^WT/WT^*, *Dsp^WT/S311A^* and *Dsp^S311A/S311A^* male mice at different ages. Bars represent s.d. Differences among groups were determined using two-way ANOVA with Šídák’s test for multiple comparisons. Each value represents the percentage of TUNEL-positive cardiomyocytes quantified in 3 different fields within the LV sub-epicardial region, from 4 non-consecutive sections per heart. Three hearts for each group were analyzed. (**H**) Confocal immunofluorescence on ventricular sections from 2 months *Dsp^WT/WT^*, *Dsp^WT/S311A^* and *Dsp^S311A/S311A^* male mice, stained with an antibody to CD8. Nuclei were counterstained with DAPI. Images are details from the LV. (**I**) Quantification, by FACS analysis, of the percentage of CD4 or CD8 positive over the fraction of CD45-positive cardiac interstitial cells. The analysis was performed in hearts from 2 months *Dsp^WT/WT^* and *Dsp^S311A/S311A^* male mice. Bars represent s.d. Differences among groups were determined using two-way ANOVA with Uncorrected Fisher’s LSD test for multiple comparisons. (Each value represents the pool of cells isolated from 2 hearts. n=6 hearts/group. *, p<0.05; ***, p<0.001).

### Contractile and electrocardiographic alterations in *Dsp^S311A^* mice

Cardiac function was assessed in male and female *Dsp^WT/WT^, Dsp^WT/S311A^* and *Dsp^S311A/S311A^*mice by echocardiography, at different time points (1, 2, 4, 6, 9 and 16 months). The very low survival of homozygous mice only allowed for comparative analyses until 9 months and the rare 12 months *Dsp^S311A/S311A^* mice were used to assess cardiac structure. While LV systolic function remained normal in the heterozygous mice until the latest time point, *Dsp^S311A/S311A^* mice showed progressive age-related decline of cardiac contractility (with respect to controls and heterozygotes), which started at 2 months and affected similarly both genders (**Fig.1C** and **Suppl.Fig.3**). In parallel, heart electrical activity was assessed by both telemetry- and surface-electrocardiography (ECG). ECG in conscious 4 months-old mice evidenced decreased heart rate (HR) in homozygotes, compared to age- and sex-matched controls, while a trend to lower HR was observed in the heterozygotes (**Suppl.Fig.4A**). While no significant differences were seen in the PR interval, the QRS duration was prolonged in both male and female homozygous and heterozygous mice of 4 and 9 months, respectively (**Fig.1D-E; Suppl.Fig.4B**). Notably, the ECG acquired in freely moving mice revealed, in both genotypes, frequent ventricular extra-systole which had sometimes a normal PR coupling, suggesting a distal conduction defect (**Fig.1F-G; Suppl.Fig.4C**). In addition, we observed trains of ventricular beats, which were frequently 3-to-10 beats long and sometimes sustained for more than 2 minutes (**Fig.1H-L; Suppl.Fig.4D**). To test the effects of adrenergic stress on arrhythmic vulnerability, we injected mice with noradrenaline while recording surface-ECG. While in control mice noradrenaline elicited the expected sinusal tachycardia, it frequently caused ventricular arrhythmias in both *Dsp^WT/S311A^* and *Dsp^S311A/S311A^* mice (**Suppl. Fig.4E**).

**Figure 3.**
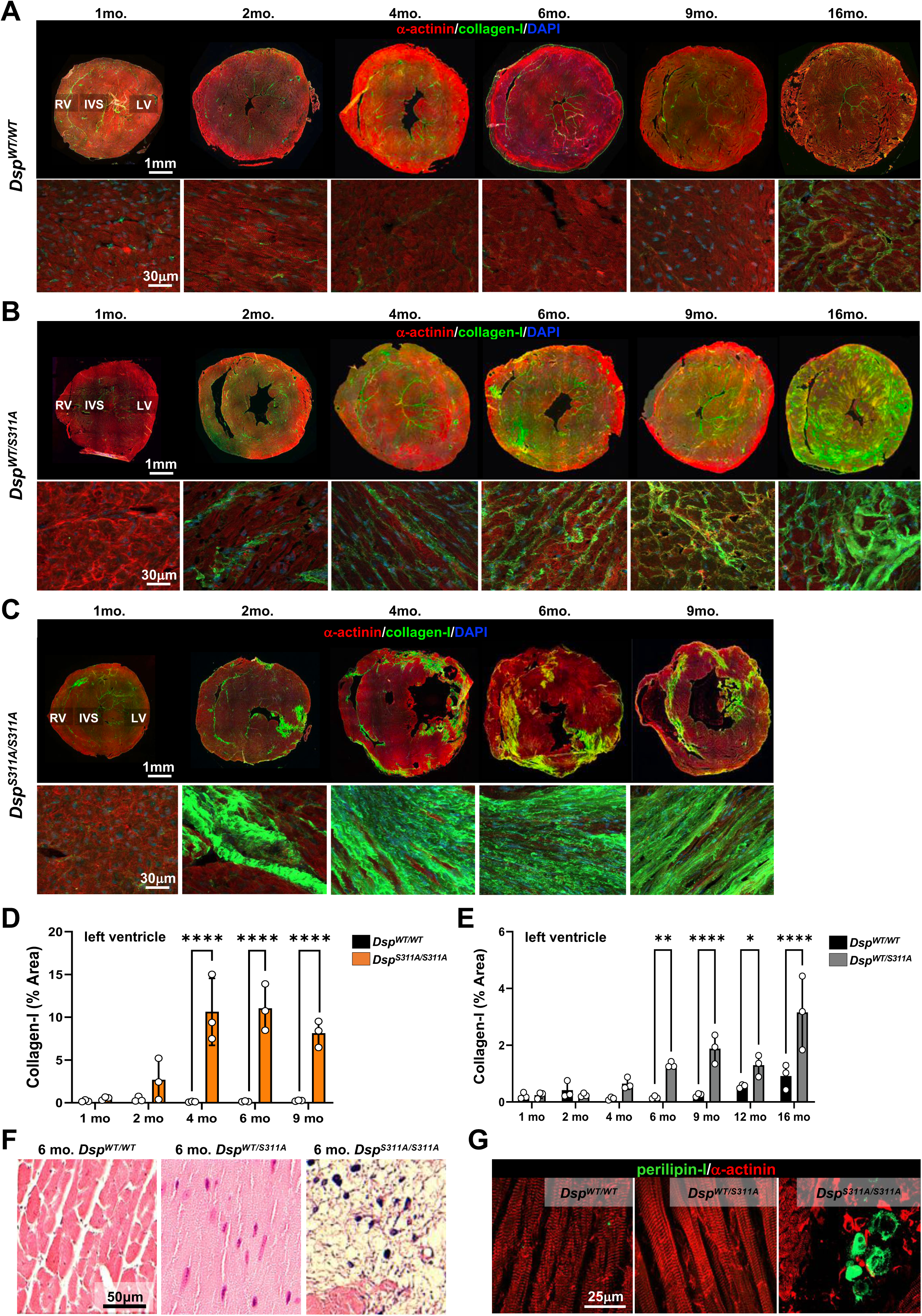
Age-dependent heart remodeling in *Dsp^S311A^* mice. (**A-C**) Confocal immunofluorescence of heart sections from the mid portion of the ventricles of *Dsp^WT/WT^* (**A**), *Dsp^WT/S311A^* (**B**) and *Dsp^S311A/S311A^* (**C**) male mice, co-stained with antibodies to collagen-I and sarcomeric actinin (α-actinin). Nuclei were counterstained with DAPI. The bottom images in (**A-C**) are details from the LV wall. (**D-E**) Quantification of the percentage of myocardial area of the LV occupied by collagen-I deposition, in heart sections from *Dsp^WT/WT^*, *Dsp^WT/S311A^* and *Dsp*^S^*^311A/S311A^* male mice at different ages. Differences among groups were determined using two-way ANOVA with uncorrected Fisher’s LSD test for multiple comparisons. (Each value represents the mean of 4 non-consecutive sections from 3 different hearts. *, p<0.05; **, p<0.01; ****, p<0.0001). (**F**) Haematoxylin-eosin staining of sections from the mid portion of the ventricles of 6 months *Dsp^WT/WT^*, *Dsp^WT/S311A^* and *Dsp^S311A/S311A^* male mice. Images are details from the LV lateral wall. (**G**) Confocal immunofluorescence on ventricular sections from 6 months *Dsp^WT/WT^*, *Dsp^WT/S311A^* and *Dsp^S311A/S311A^* male mice, co-stained with antibodies to perilipin-1 (PLIN1) and α-actinin. Images are details from the LV subepicardial region.

**Figure 4.**
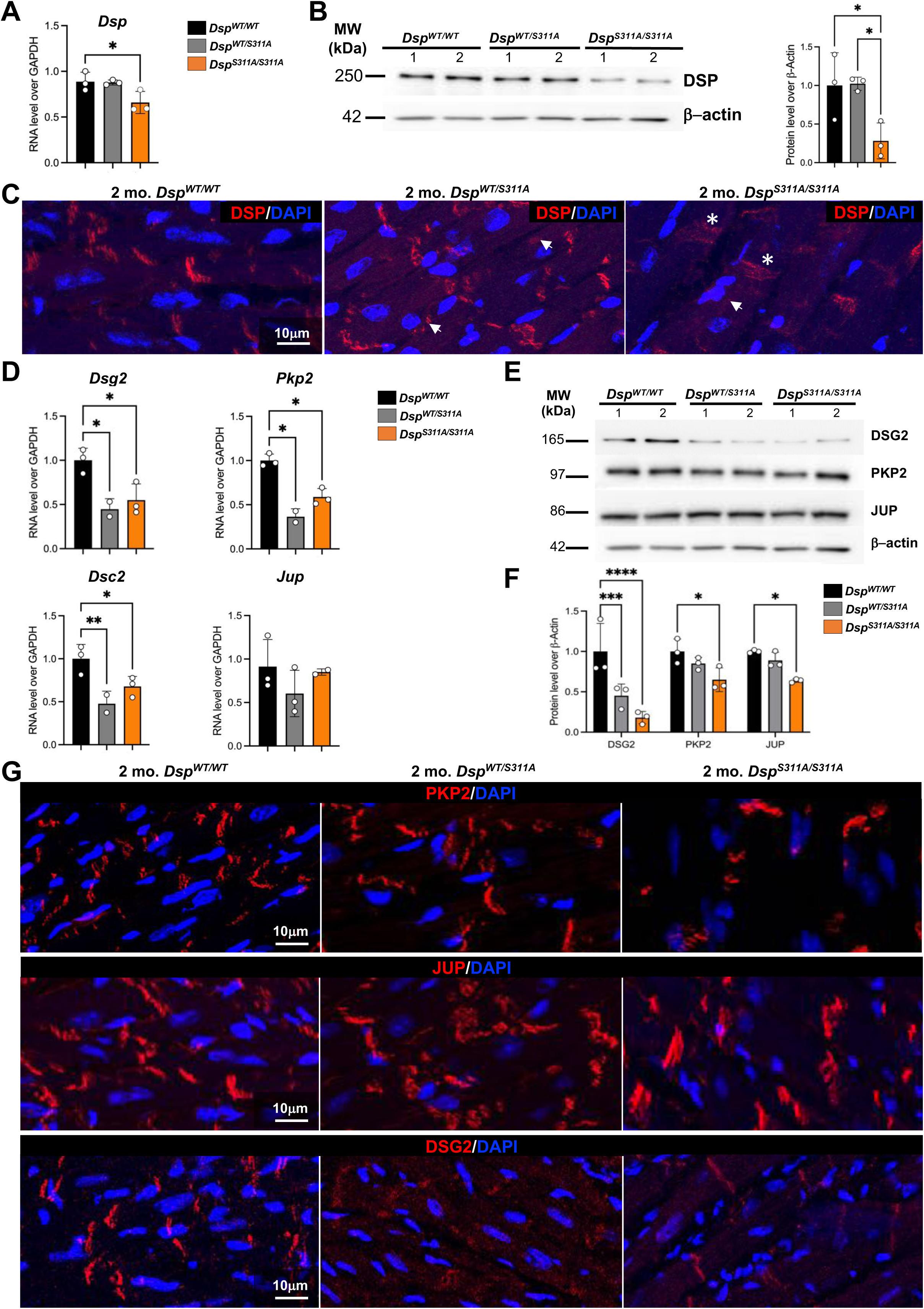
Molecular composition of desmosomes in *Dsp^S311A^* hearts. (**A**) RTqPCR on heart extracts from 2 months *Dsp^WT/WT^*, *Dsp^WT/S311A^* and *Dsp^S311A/S311A^* male mice. The expression level of *Dsp* was assessed. Bars represent s.d. Differences among groups were determined using ordinary one-way ANOVA with Dunnett’s test for multiple comparisons. (Each value represents the pool of extracts from 2 hearts. A total of 6 hearts/group was analyzed. The experiment was repeated three independent times. *, p<0.05). (**B**) Western blotting on heart extracts from 2 months *Dsp^WT/WT^*, *Dsp^WT/S311A^* and *Dsp^S311A/S311A^* male mice. Heart DSP protein content was assessed. β-actin was used to ensure equal protein loading. The graph on the right shows the relative densitometric analysis. Bars represent s.d. Differences among groups were determined using ordinary one-way ANOVA with Tukey’s test for multiple comparisons. (Each point in the graph represents the pool of extracts from 3 different hearts. A total of 9 hearts for each group has been analysed. The experiment was repeated three independent times. *, p<0.05). (**C**) Confocal immunofluorescence on ventricular sections from 2 months *Dsp^WT/WT^*, *Dsp^WT/S311A^* and *Dsp^S311A/S311A^* male mice, stained with an antibody to DSP. Nuclei were counterstained with DAPI. Images are details from the LV. (**D**) RTqPCR on heart extracts from 2 months *Dsp^WT/WT^*, *Dsp^WT/S311A^* and *Dsp^S311A/S311A^* male mice. The expression level of different desmosomal genes was assessed. Bars represent s.d. Differences among groups were determined using ordinary one-way ANOVA with Tukey’s test for multiple comparisons. (Each point in the graph represents the pool of extracts from 3 different hearts. A total of 9 hearts for each group has been analysed. The experiment was repeated three independent times. *, p<0.05; **, p<0.01). (**E**) Western blotting on heart extracts from 4 months *Dsp^WT/WT^*, *Dsp^WT/S311A^* and *Dsp^S311A/S311A^* male mice. The protein content of different desmosomal proteins was assessed. β-actin was used to ensure equal protein loading. (**F**) Densitometric analysis of western blotting in (**E**). Bars represent s.d. Differences among groups were determined using two-way ANOVA with Tukey’s test for multiple comparisons. (Each point in the graph represents the pool of extracts from 3 different hearts. A total of 9 hearts for each group has been analyzed. The experiment was repeated three independent times. *, p<0.05; ***, p<0.001; ****, p<0.0001). (**G**) Confocal immunofluorescence on ventricular sections from 2 months *Dsp^WT/WT^*, *Dsp^WT/S311A^* and *Dsp^S311A/S311A^* male mice, stained with antibodies to: Plakophilin-2 (PKP2) (top panel); Plakoglobin (JUP) (middle panel) or Desmoglein-2 (DSG2) (bottom panel). Nuclei were counterstained with DAPI. Images are details from the LV wall.

### Cardiac structural remodeling in *Dsp^S311A^* mice

Upon completion of the functional assays, we analyzed the gross heart morphology of DSP-mutants *ex vivo*. The heart weight/body weight ratio was unchanged among the different gender, age and genotype groups. At inspection, the surface of homozygous hearts appeared inhomogeneous, with patchy pale areas whose extension increased with ageing (**Fig.2A**). Consistently, histological analysis evidenced that the ventricular chambers were enlarged, with thin ventricular walls and aneurysms, and such phenotype developed during progression of the disease and reflected time-dependency of contractile dysfunction (**Fig.2B; Suppl.Fig.5**). Heart remodeling was more moderate in heterozygotes and only appeared at later time points (**Fig.2B; Suppl.Fig.5**). At more detailed pathological inspection, *Dsp^S311A/S311A^* hearts showed focal subepicardial areas of cardiomyocyte necrosis, which were predominantly found in the LV, already at early disease stage (i.e. 1-2 months) and featured an inflammatory infiltrate (**Fig.2C**). At different disease stages in homozygous (at 2 months onwards) *vs.* heterozygous (at 9 months onwards) hearts, large areas of tissue remodeling were detected in both the LV and RV. In both genotypes, myocardial remodeling became progressively more evident with ageing, although the histopathological features were profoundly different in homozygous *vs.* heterozygous mice (**Fig.2D; Suppl.Fig.6**). While in the former the large remodelled foci were characterized by the absence of cardiomyocytes and their ‘replacement’ with amorphous low eosinophilic scars, in the latter the muscle component was more preserved and surrounded by thick layers of extra-cellular matrix in a ‘patchy fibrosis’ pattern (**Fig.2D; Suppl.Fig.6**).

**Figure 5.**
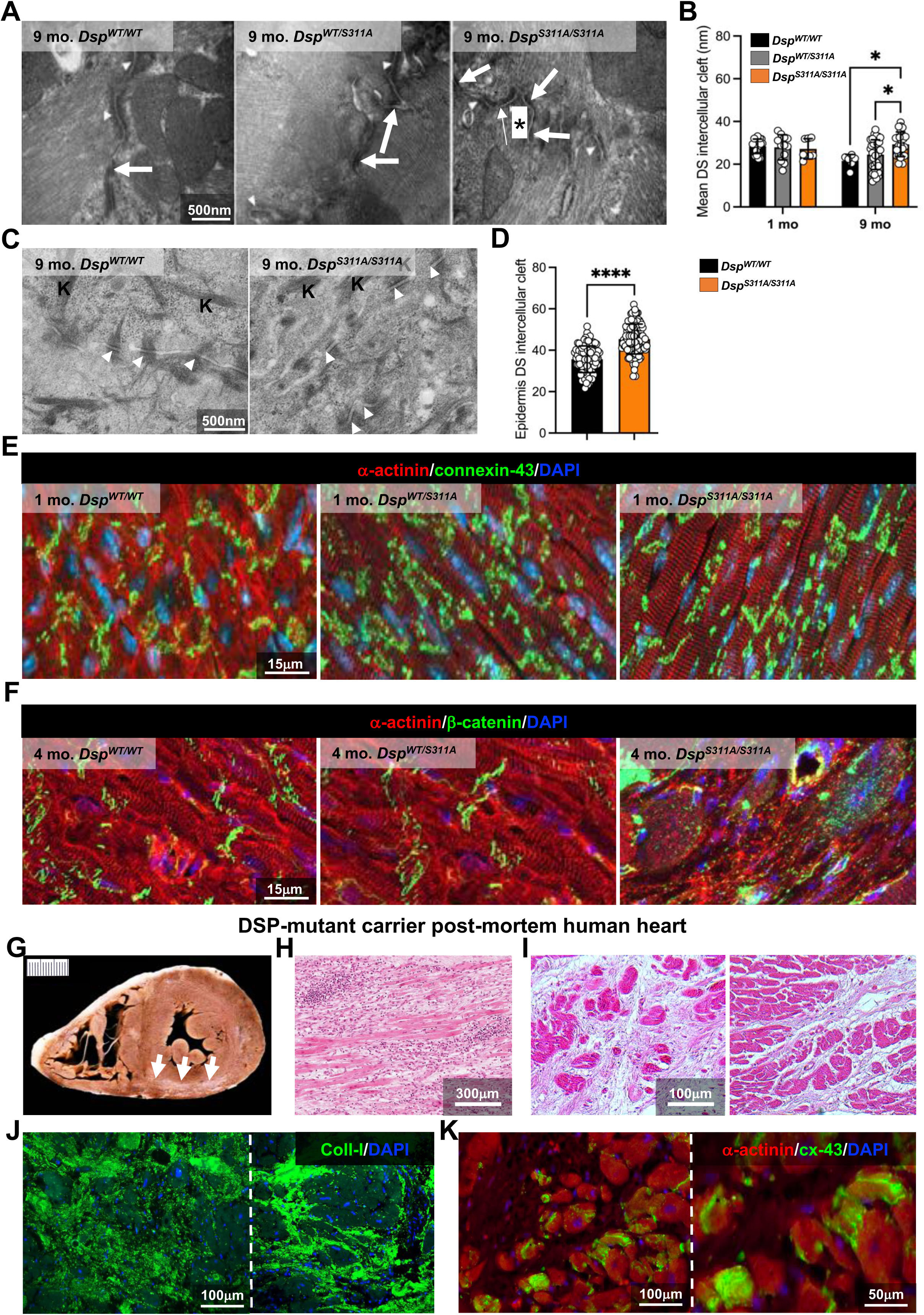
State of intercalated disks in *Dsp^S311A^* mice. (**A**) Transmission Electron Microscopy (TEM) of thin heart sections from 9 months *Dsp^WT/WT^*, *Dsp^WT/S311A^* and *Dsp^S311A/S311A^* male mice. Desmosomes (DS, arrows); Fasciae adherens (arrowheads); Repeated DS (asterisks); Gap junction (thin arrows). Images are details from the LV. (**B**) Evaluation of mean DS intercellular cleft width in 1 and 9 months *Dsp^WT/WT^*, *Dsp^WT/S311A^* and *Dsp^S311A/S311A^* male mice. Bars represent s.d. Differences among groups were determined using two-way ANOVA with Tukey’s test for multiple comparisons. (From 7 to > 37 fields from 4 different hearts/group; *, p<0.05). (**C**) TEM of thin skin sections from 9 months *Dsp^WT/WT^* and *Dsp^S311A/S311A^* male mice. (**D**) Evaluation of the mean DS intercellular cleft width in skin samples from 9 months *Dsp^WT/WT^ vs. Dsp^S311A/S311A^* male mice. Bars represent s.d. Differences among groups were determined using unpaired t-test. (n>91 fields from 4 different hearts/group; ****, p<0.0001). (**E**) Confocal immunofluorescence on ventricular sections from 1 months *Dsp^WT/WT^*, *Dsp^WT/S311A^* and *Dsp^S311A/S311A^* male mice, co-stained with antibodies to connexin-43 (cx-43) and α-actinin. Nuclei were counterstained with DAPI. Images are details from the LV. (**F**) Confocal immunofluorescence on ventricular sections from 4 months *Dsp^WT/WT^*, *Dsp^WT/S311A^* and *Dsp^S311A/S311A^* male mice, co-stained with antibodies to β-catenin and α-actinin. Nuclei were counterstained with DAPI. Images are details from the LV. (**G**) Heart from a young male individual (age range, 10-15 years) carrying the *Dsp^S299R^* variant. Arrows evidence the LV subepicardial scar. (**H-I**) Haematoxylin-eosin staining of heart sections form the heart in (**G**). Images are details from the sub-epicardial LV wall. (**J**-**K**) Confocal immunofluorescence of sections from the heart in (**G**), stained with an antibody to collagen-I (**J**) and co-stained with antibodies to α-actinin and connexin-43 (**K**). Nuclei in (**J-K**) were counterstained with DAPI.

**Figure 6.**
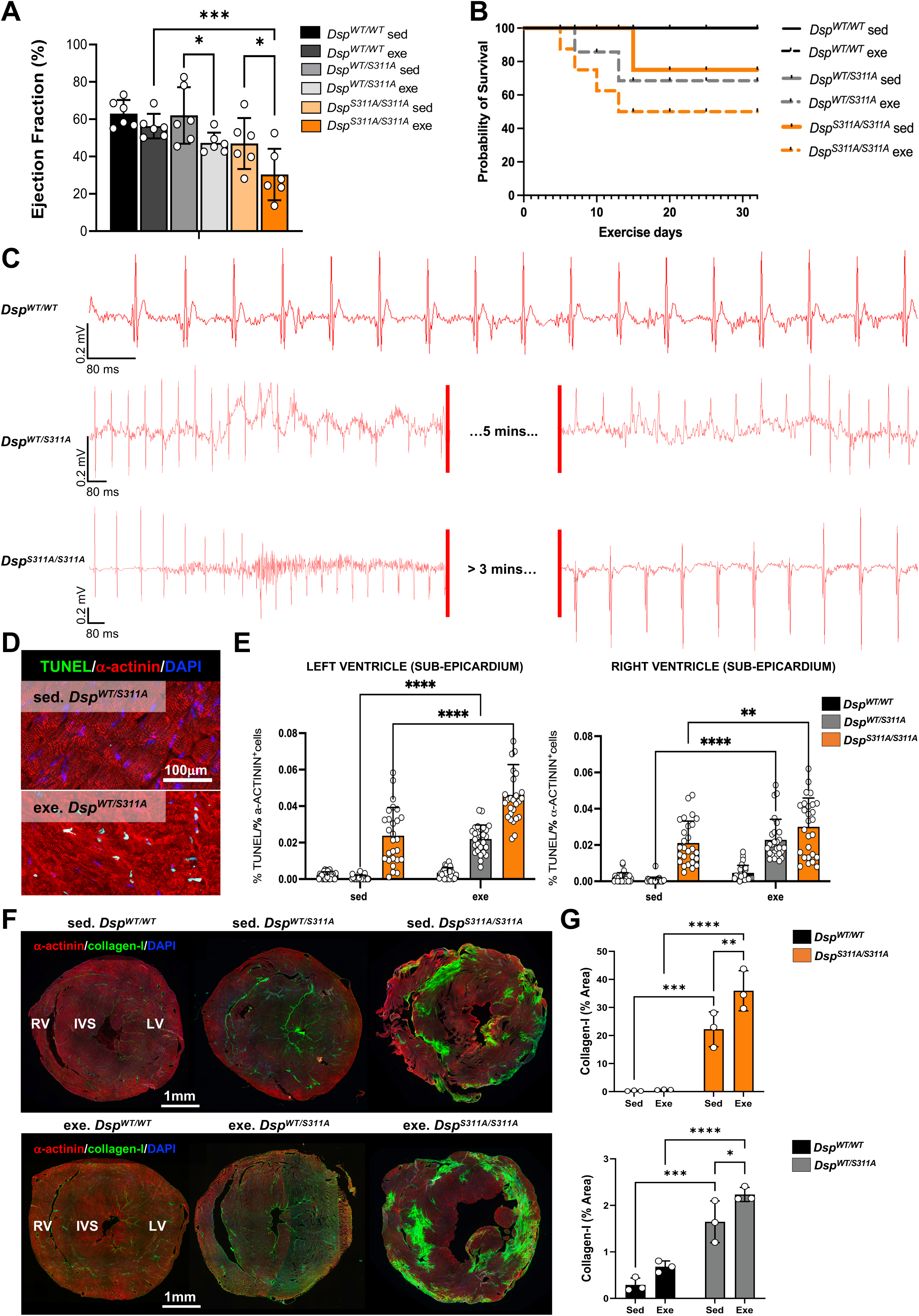
Effects of moderate exercise on cardiac phenotype of *Dsp*^S311A^ mice. (**A**) Echocardiography in sedentary *vs.* exercised 4 months *Dsp^WT/WT^, Dsp^WT/S311A^* and *Dsp^S311A/S311A^* male mice. EF, ejection fraction. Bars represent s.d. Differences among groups were determined using ordinary one-way ANOVA with uncorrected Fisher’s LSD test for multiple comparisons. (n=6 mice/group. *, p<0.05; ***, p<0.001). (**B**) Survival curves of sedentary *vs.* exercised of 4 months *Dsp^WT/WT^, Dsp^WT/S311A^* and *Dsp^S311A/S311A^* male mice. (**C**) Representative ECG traces in *Dsp^WT/WT^*, *Dsp^WT/S311A^* and *Dsp^S311A/S311A^* male mice at the end of training. (**D**) Representative images of TUNEL assay in heart sections from 4 months old sedentary (sed.) *vs.* exercised (exe.) *Dsp^WT/S311A^* male mice. Sections were co-stained with an antibody to α-actinin. Nuclei were counterstained with DAPI. Images are details from the LV. (**E**) Quantification of the number of TUNEL-positive cardiomyocytes in the LV and right ventricle of 4 months old sedentary (sed.) *vs.* exercised (exe.) *Dsp^WT/WT^*, *Dsp^WT/S311A^* and *Dsp^S311A/S311A^* male mice. Bars represent s.d. Differences among groups were determined using two-way ANOVA with Tukey’s test for multiple comparisons. (Each value represents the percentage of TUNEL-positive cardiomyocytes measured in 3 different fields within the sub-epicardial region, obtained from 4 non-consecutive sections per heart, n=3 hearts. **, p<0.01; ****, p<0.0001). (**F**) Confocal immunofluorescence on ventricular heart sections from sedentary *vs.* exercised 4 months *Dsp^WT/WT^, Dsp^WT/S311A^*, and *Dsp^S311A/S311A^* male mice. Sections were co-stained with antibodies to α-actinin and collagen-I. Nuclei were counterstained with DAPI. (**G**) Quantification of the percentage of myocardial area replaced by collagen-I deposition in ventricular heart sections from sedentary (sed.) *vs.* exercised (exe.) 4 months *Dsp^WT/WT^*, *Dsp^S311A/S311A^* and *Dsp^WT/WT^, Dsp^WT/S311A^* male mice. Bars represent s.d. Differences among groups were determined using two-way ANOVA with uncorrected Fisher’s LSD test for multiple comparisons. (Each value represents the mean of 4 non-consecutive sections from 3 different hearts. *, p<0.05; **, p<0.01; ***, p<0.001; ****, p<0.0001).

### Biventricular myocardial pathology in *Dsp*^S311A^ mice

To further characterize tissue pathology, we performed TUNEL assay to determine whether apoptosis occurs in addition to cell necrosis in *Dsp^S311A^* hearts ^19^ (see **Fig.2C**). In both homozygous and heterozygous hearts, discrete areas with elevated density of apoptotic cardiomyocytes were found in the walls of both ventricles starting from 2 and 9 months, respectively (**Fig.2E-G**; **Suppl.Fig.7**). While in homozygous hearts, the number of apoptotic cells peaked at 2 months and remained higher as compared to controls afterwards, in heterozygotes this number increased progressively during ageing (**Fig.2E-G**; **Suppl.Fig.7**). Notably, in both genotypes TUNEL-positive cardiomyocytes were predominantly found in the LV sub-epicardium. In line with the proposed role of inflammation in DSP-CM ^4,14^, confocal immunofluorescence of heart sections showed significant increase in macrophages (which were detected at all disease stages in homozygotes, and from 9 months in heterozygotes) and CD8-positive lymphocytes (which hallmarked, in homozygous hearts, the early phase of appearance of cardiac remodeling, i.e. around 2 months), as determined by both immunofluorescence and FACS-analysis of isolated cardiac interstitial cells (**Fig.2H-I; Suppl.Fig.8**). Tissue imaging and confocal immunofluorescence were then used to finely characterize the nature and composition of remodeled myocardial regions in the two genotypes. Homozygous hearts showed ‘replacement fibrosis’, revealed by anti-collagen-I staining, which appeared in discrete foci (predominantly in the LV sub-epicardium) in 2-months hearts, while larger fibrotic areas, occupying nearly half of the LV, were found in older animals. From 4-month onwards, focal collagen-I replacement was also detected in the RV wall and Interventricular Septum (IVS) (**Fig.3A, C-D; Suppl.Fig.9-12**). In addition, histological analyses and anti-perilipin-1 staining revealed small and rare fatty lesions scattered within fibrotic areas, which mainly concentrated in the LV free wall (**Fig.3F-G**). From 9 months on, we also detected areas of myocardial calcification (**Suppl.Fig.13**). In contrast, in heterozygous hearts, interstitial collagen-I enveloped cardiomyocytes within areas of ‘patchy fibrosis’, which occupied diffusely and transmurally all heart regions, and was remarkable at 16 months when heart contractility was depressed (**Fig.1C**; **Fig.3A-B, E; Suppl.Fig.9-12**).

### Molecular profile and ultrastructure of intercalated disks in *Dsp^S311A^* mice

We then used molecular biology approaches, protein biochemistry and immunofluorescence to assess the effect of the S311A-mutant allele on cardiac expression, protein content and subcellular localization of DSP. While *Dsp* gene and protein expression were comparable to controls in whole tissue extracts of heterozygous hearts, they were consistently decreased in homozygotes starting at 1 month (**Fig.4A-B**). Accordingly, confocal immunofluorescence showed that in *Dsp^S311A/S311A^* cardiomyocytes, DSP immunoreactivity was reduced and diffusely revealed at higher distance from the sarcolemma and frequently found in cytoplasmic puncta (**Fig.4C**). Interestingly, in both genotypes, the expression and protein content of the other desmosomal components were significantly lower compared to controls, with apparent subcellular distribution reflecting that of mutant DSP (**Fig.4D-G**). In line with these results, electron microscopy of intercalated discs showed the progressive deterioration of adherens junctions in *Dsp^S311A^* hearts. While in young (1 month) *Dsp^WT/S311A^* and *Dsp^S311A/S311A^* mice desmosomes and *fasciae adherentes* had mostly regular distribution with a thick electron-dense internal plaque and homogeneous intercellular clefts, in adult (9 months) AC hearts they were disarrayed (i.e. repeated desmosomes with pale internal plaques), displaying widened intercellular clefts (**Fig.5A-B; Suppl.Fig.14**). Such alterations were identified in hearts from both genders and detected focally in heterozygous and in a diffused pattern in homozygous hearts. The expression of DSP in all epidermal layers ^6,20^ and the cutaneous abnormalities found in *Dsp^S311A/S311A^* mice prompted us to analyze desmosomes in skin biopsies. Notably, skin desmosomes from both male and female adult homozygotes displayed the same abnormalities in displacement and morphometry observed in the heart (**Fig.5C-D**).

### Altered localization of Connexin-43 and β-catenin in *Dsp^S311A^* hearts

As interaction with desmosomal proteins guides the localization of gap junctions at intercalated discs ^7^ and desmosomal variants impact on β-catenin signaling ^21–23^, we addressed localization of connexin-43 and β-catenin in *Dsp^S311A/S311A^ vs. Dsp^WT/S311A^* hearts. While connexin-43 was almost entirely localized at the intercalated discs in controls, heterozygous and homozygous knock-in cardiomyocytes showed a marked delocalization of the protein with increased immunoreactivity at lateral sarcolemma and cytoplasmic aggregates (**Fig.5E**). Such cellular phenotype was detected in samples from all heart regions (i.e. LV, RV, IVS) from both male and female *Dsp* mutant mice, already at 1 month and worsened with aging (**Suppl.Fig.15-17**). In addition, β-catenin, which was found at the expected subcellular location in control hearts, was dispersed intracellularly and frequently translocated to the nucleus in both cardiomyocytes and non-cardiomyocyte cells of homozygous *Dsp^S311A/S311A^* hearts (**Fig.5F**). Consistently, *Dsp^S311A/S311A^* hearts showed increased immunoreactivity and nuclear translocation of the β-catenin co-factor LEF1 (**Suppl.Fig.18A-B**). β-catenin delocalization was not observed in heterozygous hearts, where it was localized at the intercalated disks, even at advanced disease stages (**Suppl. Fig.18C**).

### Structural remodeling in *DSP*-mutant human heart

To determine whether the histopathologic findings in *Dsp^WT/S311A^* mice replicated the abnormalities found in humans, we analyzed in detail post-mortem heart samples from a *Dsp^S299R^* variant carrier who died due to arrhythmic SD, as originally described in ^1,2^ (**Fig.5G**). Clinically, the patient presented with an incomplete right bundle branch block and mild ST segment elevation ^1,2^. Haematoxylin-eosin staining evidenced an ‘infarct-like’ band of acute–subacute myocyte necrosis associated with inflammation, in the outer mid– subepicardial layer of the postero-septal and postero-lateral LV walls (**Fig.5H-I**). The myocardium featured loose connective tissue replacement, characterized by collagen-I deposition (coexisting in areas of diffuse and patchy fibrosis), and inflammatory cell infiltrates (**Fig.5H-J**). Immunostaining also revealed delocalization of connexin-43, which was found in cytoplasmic aggregates, predominantly in cardiomyocytes close to the remodeled myocardial areas (**Fig.5K**).

### Exercise as phenotypic modifier in *Dsp*^S311A^ mice

Based on the well-appreciated correlation between physical stress and progression of myocardial remodeling in AC, we assessed the effect of 1 hour running bouts, repeated daily for four consecutive weeks, on the histopathological and functional cardiac phenotype of *Dsp* mutant mice (see **Suppl. Methods**). As expected, exercise impacted on disease progression and, at the end of the protocol (corresponding to 4 months of age), *Dsp^WT/S311A^* and *Dsp^S311A/S311A^* mice had a significant decline in cardiac function, compared to isogenic age-matched sedentary controls (**Fig.6A**). Notably, as shown in (**Fig.6B**), a large fraction of exercised mice, from either strain, died during the period of exercise protocol, frequently due to cardiac arrest occurred shortly after completion of one of the bouts. Whilst we were unable to record lethal arrhythmic events, complex sustained arrhythmic episodes were frequently detected in both genotypes during the period of exercise (**Fig.6C; Suppl.Fig.19A**). Histopathological analyses of hearts harvested after exercise showed that, irrespective of the animal gender, exercise accelerated myocardial remodeling in both genotypes. Indeed, in both *Dsp^WT/S311A^* and *Dsp^S311A/S311A^* mice the percentage of TUNEL-positive cardiomyocytes, which were found largely in the LV sub-epicardium, increased compared to non-exercised animals (**Fig.6D-E; Suppl.Fig.19B-C**). This was accompanied by increased fibrotic remodeling, as shown by the enhanced collagen-I deposition (**Fig.6F-G**; **Suppl.Fig.19D-E**).

## DISCUSSION

We here characterized the cardiac phenotype of a novel knock-in mouse strain, modelling the recessive- and dominant-forms of *DSP*-linked ‘arrhythmogenic’ diseases, namely i.e. the cardio-cutaneous Carvajal syndrome and DSP-CM, respectively. Consistent with a recessive inheritance pattern, homozygous *Dsp^S311A^* mutant mice exhibit dilated cardiomyopathy with significant LV involvement and systolic dysfunction, besides arrhythmic SD, along with characteristic skin abnormalities. *Dsp^WT/S311A^* mice recapitulate key clinical features of DSP-CM, a left dominant variant of AC with elevated arrhythmogenic vulnerability and fibrotic remodelling disproportionate to contractile dysfunction.

Arrhythmogenic Cardiomyopathy is a rare familial cardiac disorder responsible for a significant proportion of stress-related arrhythmic SD in the young and athletes ^11–16,19,21–27^. In most cases, AC is caused by mutations in genes encoding desmosomal proteins (i.e. *DSP, DSG2, DSC2, PKP2, JUP*), which form desmosomes connecting adjacent cardiomyocytes ^11–16,19,21–36^. Although most patients express AC-variants in heterozygosity, recessively inherited cases often linked to skin/hair abnormalities, as seen in Naxos and Carvajal diseases, have been reported ^8–10,32^. AC hearts feature cardiomyocyte death, tissue inflammation, and myocardial fibro-fatty replacement, which affect both electrophysiology and contractility, favoring respectively arrhythmias and heart failure ^11–16,19,21–39^. Variants in *DSP* were initially identified in patients with the autosomal recessive cardio-cutaneous Carvajal syndrome ^8–10^. Several dominant-inheritance *DSP* variants were since recognized, clustering with left-dominant AC, which present with high incidence of PVCs and LV fibrotic remodeling (often without RV involvement), frequently preceding LV dysfunction and dilation ^1–5,14,15,40,41^. Notably, episodes of acute myocardial injury are common and are accompanied by myocardial inflammation, which has been proposed to have a mechanistic role in the disease pathogenesis ^2–4,14,15^. Unlike the classic right-dominant form of AC, traditional diagnostic criteria, including those used in sport pre-participation screening, are ineffective in DSP-CM and risk-stratification and prediction remains a challenge ^13–15,39–42^.

Structurally, DSP is a tripartite plakin-family cyto-linker that bridges transmembrane cadherins and desmosomal armadillo proteins to intermediate filaments (i.e. desmin in cardiomyocytes) through a central rod domain that enables DSP dimerization ^43^. Beyond its structural role in desmosomes, the multiple functions of DSP ^6,43–46^ (see Introduction) explain the embryonic lethality of DSP-null mice ^46^, and the effects of its pathogenic variants on the cutaneous phenotypes (i.e. striate palmoplantar keratoderma and lethal acantholytic epidermolysis bullosa), which may associate or not with cardiac abnormalities, i.e. the Carvajal syndrome ^8–10,47,48^. Several missense and non-missense variants of *DSP* gene have been identified, which lead to either truncated- or point mutated-proteins ^4^. While it is generally thought that the former operate through protein haploinsufficiency, and the latter through dominant-negative effects, the ‘genotype-clinical phenotype’ correlation is still inconclusive ^4,49^. While the high arrhythmogenic risk is associated to all *DSP* variants, LV dysfunction mainly occurs in carriers of truncated DSP, and was attributed to disrupted interaction with intermediate filaments (as in the Carvajal syndrome) ^1,4,10,49^. In parallel, the variable phenotypes of missense variant carriers were suggested to reflect the peculiar mutation site, which reappraised the value of a genotype-specific approach for diagnosis and risk stratification in DSP-CM ^4^. The recognition that both cardiac and extra-cardiac cells harboring *DSP*-variant may contribute to the disease pathogenesis supports that DSP-CM is a multicellular ailment, rather than a cardiomyocyte-restricted disease ^50–52^. This factor is disregarded in the different transgenic models with cardiomyocyte-specific expression of the disease-genetics, each replicating individual aspects of myocardial remodeling (i.e. cardiomyocyte apoptosis, fibrosis, ultrastructural abnormalities of intercalated discs, ventricular dilation and contractile dysfunction), while understating the intercellular and interorgan effects within variant-bearing organism ^23,33,34,53–58^. For a comprehensive study of DSP-CM, we generated a pan knock-in mouse model carrying a pathogenic variant of the *Dsp* gene. Among the missense mutation sites described up to now, we focused on S299, which is located in the predicted Plakoglobin-interaction domain at the N-terminal portion and is critical for DSP incorporation into desmosomes and their integrity ^1,2,59,60^. In addition, S299 is the only PKC (Protein Kinase C) phosphorylation site conserved in the N-terminal of all PKC-regulated plakins, which may further impact on DSP interactions with desmosomal components at the plasma membrane ^1^. We initially aimed at reproducing the human Ser-to-Arg substitution, by editing the murine ortholog S311 (*Dsp^S311R^*). Bioinformatic prediction of protein structure indicated that Ser substitution with the positively charged Arg would interfere with the interaction between the -OH group of Ser and the nearby Asp309 and Asn464, leading to a dramatic destabilization of the protein N-terminal domain. Consistently, this single amino acid replacement had the same effect of global DSP knock-out, as homozygous mice died in utero at P4. We therefore replaced Ser-311 with Ala which was predicted to only slightly destabilize the protein structure, while ablating the PKC-dependent phosphorylation site. These *Dsp^S311A^* knock-in mice were viable, fertile and born at the expected Mendelian ratio, thus allowing deeper characterization of the cardiac phenotype. Remarkably, *Dsp^S311A^* transgenics bred into hetero- or homo-zygous genotypes developed in time distinct clinical/pathological phenotypes.

Surprisingly, homozygous mice replicated the disease phenotype of the cardio cutaneous Carvajal syndrome, which was up to now associated to non-missense variants at C-term leading to truncated DSP ^1,2,8–10,49^. Indeed, *Dsp^S311A/S311A^* mice showed a dilatative cardiomyopathy with extensive scarring, ventricular aneurysms and cutaneous defects (i.e. alopecia) which, as in patients, manifested at young age. Hearts showed LV sub-epicardial areas of cardiomyocyte necrosis and apoptosis, infiltrated by CD8-positive cells and macrophages, which were also detected at advanced ages. Mice developed precocious contractile dysfunction, which progressed during ageing and resulted in heart failure before 6 months. Such contractile dysfunction was accompanied by myocardial remodelling, which evolved in time with diffuse replacement fibrosis (from 2 months onwards), focal fatty tissue (from 6 months) and tissue calcification (at 9 months), which culminated in thinning of the LV free wall and chamber dilatation. Although the LV was mainly affected, the RV was variably involved. Notably, *Dsp^S311A/S311A^* mice showed a significant increase in the number of PVCs with distal conduction defect and sustained arrhythmias, which may explain the increased incidence of SD observed at rest from early disease stage (i.e. 4 months) on. Differently from the homozygotes, *Dsp^WT/S311A^* mice did not show evident cutaneous alterations, had normal lifespan and contractile dysfunction appeared only later (from 16 months). Telemetry-ECG revealed frequent arrhythmic events already under unstressed conditions, closely mirroring patients’ phenotypes, including QRS prolongation, frequent isolated PVCs, sustained monomorphic ventricular tachycardia and distal conduction defects. Notably, as observed in certain patient subgroups, arrhythmic episodes in these mice occurred earlier than, and independently of contractile dysfunction ^1,2,61^. This early electrical instability, preceding structural pathology, aligns with often-reported discrepancy between arrhythmic vulnerability and structural remodeling, in DSP-mutant patient cohorts ^1–5,14,60,61^. Our findings, obtained in both murine and patient hearts, suggest that the electrical dysfunction may be due to displacement of connexin-43 from cardiomyocyte intercalated disks, which was observed in both hetero- and homo-zygous hearts as early as 1 month of age, and notably in heterozygotes even in the absence of structural myocardial remodeling. Fibrotic remodeling, in heterozygous myocardium, developed later than in homozygous hearts, and was associated to increased cell death and inflammatory response. This led to elevated collagen-I deposition which expanded interstitial spaces between cardiomyocytes and extended from the LV to all heart regions with age and paralleled with progressive contractile dysfunction. Remarkably, pathology in this mouse model was not associated to gender, consistent with clinical observation in DSP-CM patients ^1–5,15^. The comparative analyses of the two genotypes prompted us to speculate on the correlation between protein variant and its effect. While in heterozygotes cardiac DSP expression and protein content were normal, in homozygous hearts the sole point variant was sufficient to reduce DSP expression and protein content to levels comparable to that of haploinsufficiency. Remarkably, in both genotypes, expression of the *Dsp^S311A^* variant, which likely affects protein-protein interaction at plasma membrane (i.e. through altered DSP conformation or ablation of its putative phosphorylation site), causes reduction of almost all the other desmosomal components at both mRNA and protein levels. This data is in line with the results obtained in other models of AC-linked variants ^62^, suggesting possible transcriptional roles of these proteins and that formation of stable desmosome is required for protein stability. Notably, such mechanism links a missense mutation in a single desmosomal component to its effect of the molecular structure of the entire multiprotein complex, which may reflect on the aggressiveness of specific disease genotypes.

Further insight into DSP-CM pathogenesis may arise from investigating the effects of moderate chronic exercise on disease progression and manifestations. While the link between acute adrenergic stress (i.e. acute noradrenaline injection) and increased incidence of arrhythmias is well accepted in several familial arrhythmogenic disorders, the role of exercise in DSP-CM remains debated ^1,2,63,64^. Our findings indicate that moderate chronic exercise acts as a negative modifier of DSP-CM, as it heightened arrhythmic events, accelerated structural remodeling and reduced survival in both mouse genotypes.

Taken altogether, our results demonstrate that *Dsp^S311A^* knock-in mice well resemble several clinical aspects described for hetero- and homo-zygous forms of cardiomyopathies linked to a missense *DSP*-variant. Our novel knock-in mice are suited to study disease mechanisms in Carvajal syndrome and DSP-CM and answer relevant questions yet unaddressed, regarding, as example, the mechanisms underlying cell death, the role of inflammation and the involvement of extra-cardiac cell populations. In addition, the *Dsp^S311A^* murine strains may be adequate to cell and molecular biological studies to uncover desmosome-unrelated roles of DSP and the mechanistic insight in cross-regulation of desmosomal proteins. More in general, *Dsp^S311A^* knock-in mice represent unique preclinical models to test therapeutic interventions aimed at interfering with specific ‘time-dependent’ pathological aspects of the disease.

## Data Availability

Data will be made available upon manuscript publication

## ABBREVIATION LIST

AC: Arrhythmogenic Cardiomyopathy
DSC-2: DeSmoCollin-2
DSG-2: DeSmoGlein-2
DSP: DeSmoPlakin
DSP-CM: DeSmoPlakin CardioMyopathy
ECG: ElectroCardioGraphy
HR: Heart Rate
IVS: InterVentricular Septum
JUP: plakoglobin
LV: Left Ventricle
PKP-2: PlaKoPhilin-2
PVC: Premature Ventricular Contraction
RV: Right Ventricle
SD: Sudden Death
TEM: Transmission Electron Microscopy
WT: Wild Type

## FUNDING

This work was supported by Italian Ministerial grants PRIN 2022F3NENH to **M.M.**, PRIN 20229PX74A to **T.Z.** and PRIN 202249XEA5 to **C.B**. **C.B., S.R.** and **M.D.B.** are supported by the registry for Cardio-Cerebro-Vascular Pathology, Veneto Region, Italy. **A.G.** is supported by the framework of the National Recovery and Resilience Plan (NRRP), Mission 4, Component 2, Investment 1.4, funded by the European Union - Next Generation EU, Project CN00000041, CUP C93C22002780006, Spoke n. 4 "Metabolic and Cardiovascular diseases". **I.P.V** is supported by the European Union’s Horizon 2020 research and innovation programme under the Marie Skłodowska-Curie grant agreement No 101034319 and from the European Union – NextGenerationEU. **J.M.P.P.** is supported by grants PID2021-122626B-I00 and RED2022-134485-T from the Spanish Ministry of Science and Innovation-AEI. **M.L-M.** is the recipient of a national PhD training fellowship (PRE2022-104033).

## DISCLOSURES

“The authors have declared that no conflict of interest exists”

## AUTHOR CONTRIBUTION

**A.D.B.** generated *Dsp*^S311A^ knock-in mice and performed initial experiments of functional and structural cardiac phenotyping; **A.G.** and **I.P.V.** characterized cardiac functional and structural phenotype, performed statistical analysis, and contributed to manuscript preparation; **R.B.** performed echocardiography, analysed data and contributed to manuscript preparation; **M.C.D.S.** performed confocal immunofluorescence analyses; **M.A.** performed FACS analysis; **N.K**. and **P.D.** generated *Dsp*^S311A^ knock-in mice; **V-D.M.** critically discussed data; **R.C.** contributed to genome sequencing analysis and mouse genotyping; **A.S.** analysed ECG dat**a; M.L-M.** performed immunofluorescence analyses; **M.D.G.** analyzed human samples; **M.D.B.** performed ultrastructural analyses; **S.R.** performed histological analyses on heart samples; **D.C.** critically discussed data; **B.B.** critically discussed ECHO and ECG data; **G.Z.** performed the 3-D reconstruction of mutated protein; **G.T.** critically discussed data, revised and contributed to manuscript preparation; **K.P.** contributed to genetic analyses, critically discussed data and contributed to manuscript preparation; **J.M.P.P.** supervised some immunofluorescence analyses, critically discussed data and contribute to manuscript preparation; **M.P.** supported and supervised the generation of knock-in mice; **C.B.** contributed to study design, critically discussed data and contributed to manuscript preparation; **M.M**. and **T.Z.** designed the study, supervised experiments, analyzed data and wrote the manuscript. All authors approved the final version of the manuscript and agree to be accountable for all aspects of the work, ensure all parts of the work are appropriately investigated and resolved. All persons designated as authors qualify for authorship and have been listed.

## Notes

### Competing Interest Statement

The authors have declared no competing interest.

### Author Declarations

Human samples: We analysed a heart from a young male individual (age range 10-15 years) carrying the DspS299R variant and suddenly died at rest. The heart was acquired during routine post‐mortem clinical investigations and archived at the Cardiovascular Pathology Unit of the University Hospital of Padova. The sample was used in accordance with the 'Recommendation (CM/Rec (2016) 6) of the Committee of Ministers of the Member States on research on biological materials of human origin', released by the European Council as received by the Italian National Council of Bioethics which reviewed and confirmed adherence to the principles outlined in the recommendation, ensuring that the dignity, privacy, and rights of individuals were fully protected.

