## Supplementary Data for "*Dsp^S311A^* knock-in mice replicate the clinical-pathological features of dominant and recessive forms of Desmoplakin-related cardiomyopathies"

5, Structure Fédérative de Recherche Necker, INSERM US24/CNRS UAR 3633, Paris, France.

6, Institut Imagine, Paris, France;

7, Department of Animal Biology, University of Malaga & Instituto de Biomedicina de Malaga-Plataforma BIONAND, Spain;

8, Université Paris Cité, CNRS, INSERM, Institut Necker Enfants Malades-INEM, Paris, France.

***, equal contribution**

**Short title: Novel murine model of Desmoplakin-Cardiomyopathy**

**Conflict-of-interest statement:** "The authors have declared that no conflict of interest exists"

**Correspondence to:**

**Tania Zaglia, PhD,**

Department of Cardiac, Thoracic, Vascular Sciences and Public Health, University of Padova, via Giustiniani 2, 35128 Padova, Italy

Department of Biomedical Science, University of Padova, via Ugo Bassi 58/B, 35131 Padova, Italy

#### Supplementary Data content:

**-Supplementary Methods;**

**-Supplementary Figures (1-19);**

**-Supplementary Figure Legends (1-19);**

**-Supplementary Tables (1-2);**

**-Supplementary References.**

#### Supplementary Methods

**Guide design and cloning.** RNA guides were designed by manual inspection of the mouse genomic sequence flanking serine 311 of the *Dsp* gene and evaluated for potential off-target activity using CRISPOR (http://crispor.gi.ucsc.edu/). The following oligonucleotides (5' to 3') were ordered from Eurofins and annealed for cloning.

| | |
|---|---|
| **sense** | caccG<u>AGGAGGAGCTGCTCTATGAC</u> |
| **antisense** | aaacGTCATAGAGCAGCTCCTCCTC |

The chosen guide sequence (underlined) complements the *Dsp* gene in exon 7 and is within 3 nucleotides of the desired variant site for increased knock-in efficiency (**Suppl.Fig.1B**). An extra G was added in for proper transcription initiation from the U6 promoter. The lowercase bases are the overhangs needed for cloning. The oligonucleotide pair was ordered from Eurofins and annealed before ligation into pX459, a kind gift from Feng Zhang (#62988, Addgene) and that expresses Cas9 and puromycin resistance along with a U6-driven expression of the guide RNA using the Zhang lab recommended protocol as provided by Addgene.

**Expression of the CRISPR-Cas9 components in mammalian cells.** To validate the gRNA in DNA cutting, NIH 3T3 cells were transfected with the pX459-targeting construct, and transfected cells were enriched by selection with puromycin (1 μg/ml) for 48 hours.

Single clones were then isolated and allowed to grow in 96-well plates before analysis by Sanger sequencing to determine cutting efficiency. Data indicated that the guides and Cas9 worked (over 30% targeting efficiency), and we thus progressed to design the repair template.

**Repair template**. The single-stranded DNA oligonucleotide repair template (ssODN) was synthesized by IDT and contains 50nt homology arms flanking the targeted variant site. The ssODN also contains 4 silent variants (underlined) to prevent re-cutting and also allow for rapid genotype screening using the restriction enzymes XmnI and RsaI (in blue and green respectively).

| | |
|---|---|
| **ALA** | 5'- ATC CAG GCC ACC TCT CGA GAG ATC ATG TGG ATC AAT GAC TGC GAG GAG GAA GAG CTT CTG TAC GAC TGG GCC GAC AAG AAC ACC AAC ATC GCT CAG AAG CAG GAG GCC TTC TCT GTA AGA TG -3' |

**Cell transfection with the spCas9, gRNA and repair template.** NIH 3T3 (ATCC® CRL-1658™) cells were seeded onto 6-well plates at a concentration of 1 x $10^5$ cells/well and maintained in standard Dulbecco's modified Eagle's medium (DMEM, Life technologies), supplemented with 10% FBS (Gibco, ThermoFisher) and 1% P/S (Gibco, ThermoFisher). One $\mu$g of the pX459_gRNA and 100pmol of the ssODN were transfected using Lipofectamine 3000 (ThermoFisher). Forty-eight hours after transfection, the cells were trypsinized and plated into 15 cm dishes in medium containing 1ug/ml puromycin. After another 48 hours, the medium was replaced with fresh medium without puromycin. After cells began to grow clonally, they were picked by scraping and added to individual wells of a 96-well plate. Each subclone was then analyzed by Sanger sequencing.

**Mouse genotyping.** Mouse genotyping was performed using the Rapid Extract PCR Kit (PB10.24-40) from PCR Biosystems, followed by amplification of the target region using the

HS Taq Mix Red according to the manufacturer's instructions. The amplified product was then processed for Sanger sequencing [65] (see **Suppl. Table 1**).

**Whole genome sequencing.** Genomic DNA was extracted from approximately 10mg of snap frozen heart tissue from knock-in mice, by using the Nucleic acid isolation kit I (Roche Applied Science, Mannheim, Germany). For tissue lysis, 200µl of MagNA Pure Compact DNA Lysis buffer (Roche Applied Science, Mannheim, Germany) were added to the tissue sample, which was processed with a Tissue Lyser System (Qiagen, Hilden, Germany) for 2 minutes at 25Hz. The homogenized sample was incubated with 20µl of Proteinase K (20 mg/ml; Roche Diagnostics GmbH, Mannheim, Germany) for 10 minutes at 56°C and transferred on the MagNA Pure Compact workstation. Whole genome sequencing (WGS) was performed by Biodiversa s.r.l. (Rovereto (TN), Italy), using SureSelect$^{QXT}$ Whole Genome Library Prep for Illumina Multiplexed Sequencing**.**

**Moderate chronic exercise.** Three months old normal controls ($Dsp^{WT/WT}$), $Dsp^{WT/S311A}$ and $Dsp^{S311A/S311A}$ mice underwent moderate exercise on Treadmill (Panlab, Harvard Apparatus), 1 hour/day, 5 days/week for 4 consecutive weeks. The maximum treadmill speed was progressively increased from 16 cm/s to 20 cm/s during the training period.

**Echocardiography.** Echocardiographic analysis was performed in $Dsp^{WT/S311A}$ and $Dsp^{S311A/S311A}$ mice, and littermate controls, at different ages, using a Vevo 2100™ (VisualSonics, Toronto, Canada) system, equipped with a 30 MHz transducer, following the procedure previously described by Zaglia et al. [66]

**Surface electrocardiographic (ECG) recording.** Mice were anesthetized with isofluorane (2.5-3%, v:v in $O_2$) and standard lead II ECG recording (Powerlab 8/30, Bioamp; AD Instruments) was performed. Standard ECG parameters (i.e. QRS duration, PR interval and heart rate) were calculated using the software LabChart 7.1 (AD Instruments), both at baseline and upon noradrenaline (NA, 2mg/kg, i.p.) administration.

**Telemetry-ECG.** Telemeters implantation (DataScience International, DSI PhysioTel – ETA - F10 Implant), ECG recording in conscious mice and data analysis were performed as described in Zaglia et al. [66]

**Isolation of cardiac interstitial cells and FACS analysis.** Single cells suspension was obtained by digesting mouse heart with 2mg/ml Type-IV Collagenase (Worthington Biochemical Corp, NJ, USA) and filtered through a 70µm cell strainer. Cell suspensions were incubated with Fc-Block™ (BD Biosciences) for 15 minutes on ice and then labelled with specific antibodies: anti-mouse CD45 PerCP/Cyanine5.5 (Biolegend®, CA, USA); anti-mouse/human CD11b APC/Cy7 (Biolegend®); anti-mouse Ly-6G APC (Biolegend®); anti-mouse Ly-6C PE (Biolegend®); anti-mouse CD4 PE (BD Biosciences); anti-mouse CD8 eFluor® 450 (Invitrogen). Data were acquired with a FACSCanto (BD Biosciences) cytometer and analysed using FlowJo software (BD Biosciences).

**Mouse heart harvesting and processing.** Hearts were harvested from $Dsp^{WT/WT}$, $Dsp^{WT/S311A}$ and $Dsp^{S311A/S311A}$ mice, at different ages. The apical portion of the hearts was immediately frozen in liquid nitrogen and used for molecular/biochemical analyses, while the basal portions underwent fixation, as described in Zaglia et al. [17,66], and used for immunofluorescence and morphometric analyses.

**Immunofluorescence analysis on heart cryosections.** Murine heart cryosections were processed for immunofluorescence as described in Zaglia et al. [67]. The primary antibodies used in this study are listed in **Suppl. Table 2**. Slices were analyzed at the confocal microscope (Zeiss LSM900).

**Electron microscopy analysis.** In a subset of experiments, the apical portions of the hearts from 1- and 9-month-old $Dsp^{WT/WT}$, $Dsp^{WT/S311A}$ and $Dsp^{S311A/S311A}$ mice were fixed in Karnovsky solution (2.5% glutaraldehyde, *plus* 2% paraformaldehyde) in 0.1M phosphate buffer (pH 7.2) and post-fixed in 1% osmium tetroxide for 1 h. Samples were then dehydrated in ethanol and embedded in epoxy resin, as described in Basso et al. [18]. After evaluation at light microscope of semithin sections, ultrathin sections were prepared. After staining with uranyl acetate and lead citrate, ultrathin sections were examined with Hitachi H-7000 (Hitachi, Tokyo, Japan) transmission electron microscope.

**Quantification of myocardial fibrosis.** Four non-consecutive cryosections from the mid-portion of the ventricles were stained with a collagen-I antibody. Hearts from controls and knock-in mice were analyzed at different ages. Composite images of the whole heart were obtained using a Leica microscope (Leica DM6B) and analyzed with the open-source software Fiji [68] to quantify the percentage of myocardial area occupied by collagen-I in the LV, IVS and RV.

**TUNEL assay**. This assay was performed using the DeadEnd™ Fluorometric TUNEL System Kit (#G3250, Promega), following the manufacturer's instructions. Sections were analyzed at the Zeiss LSM900 upright confocal microscope. We acquired images of 6 randomly chosen fields from both the RV and LV sub-epicardial region from 4 non-consecutive cryosections from the mid portion of the ventricles. Images were used to calculate the percentage of TUNEL positive cells over the percentage of $\alpha$-actinin positive cells.

**RTqPCR.** The analysis was performed following the protocol described in Zaglia et al. [66]. Primers used in this study are listed in **Suppl. Table 2.**

**Western Blotting.** This analysis was performed as described in Zaglia et al. [66]. Antibodies used in this study are listed in **Suppl. Table 1**.

**Statistics.** Statistical analysis was performed using GraphPad Prism 8. Normality of data distribution was assessed with Shapiro-Wilk test. In datasets showing normal distribution, unpaired *t* test with Welch's correction (comparison between two groups) or one-way analysis of variance (ANOVA) (comparison between three or more groups) were applied. For one-way ANOVA, depending on the result of SD equality (by a Brown–Forsythe test), ordinary or Brown–Forsythe and Welch ANOVA tests were performed. In cases where two factors were analyzed simultaneously (e.g. genotype and time points), a two-way ANOVA was used, followed by Tukey's, Dunnett's, Šídák's or Uncorrected Fisher's LSD test for multiple comparisons as appropriate. In non-normally distributed datasets, non-parametric tests were used. All data are expressed as the mean ± standard deviation (s.d.). Confidence intervals were set at 95% for statistical comparisons. A P value <0.05 was considered statistically significant.

#### Supplementary Figures

**Supplementary Figure 1. Generation of *Dsp*$^{S311A}$ mice.**

(**A**) Partial primary protein sequence of the human and murine Desmoplakin gene with the key conserved residues in bold. (**B**) Identification of the guide sequence (grey shaded) within 3 nucleotides of the desired variant site (in blue) in exon 7 of the mouse *Dsp* locus. (**C**) The repair template introduces four silent variants to prevent Cas9 binding and re-cutting. Silent variants for the two restriction enzymes and variants encoding the serine-to-alanine substitution are highlighted in red. (**D**) Sanger sequencing traces from *Dsp*$^{WT/WT}$, *Dsp*$^{WT/S311A}$ and *Dsp*$^{S311A/S311A}$ mice. The RNA guide sequence is represented in green, PAM sequence in red and nucleotide triplets encoding for Serine (S) and Alanine (A) in blue.

**Supplementary Figure 2. Whole genome sequencing of *Dsp*$^{S311A}$ mice.**

(**A**) Coding-variant distribution in *Dsp*$^{S311A/S311A}$ mice. The characterization of biological effects of 200000 single-nucleotide variations and small indels identified through whole genome sequencing was carried out using the Ensembl Variant Effect Predictor. (**B**) Expression levels of genes carrying large insertions/deletions among different tissues using the EMBL Expression Atlas. Dark and light blue indicates medium and low expression, respectively. (**C**) Table of genes having insertions/deletions larger than 20 nucleotides.

**Supplementary Figure 3. Echocardiographic assessment of *Dsp*$^{S311A}$ mice.**

(**A**) Representative images of echocardiographic analysis on 4 months *Dsp*$^{WT/WT}$, *Dsp*$^{WT/S311A}$ and *Dsp*$^{S311A/S311A}$ male mice. (**B**) Evaluation of Ejection Fraction (EF) and Fractional Shortening (FS) in *Dsp*$^{WT/WT}$, *Dsp*$^{WT/S311A}$ and *Dsp*$^{S311A/S311A}$ male mice at different ages. Differences among groups were determined using two-way ANOVA with Tukey's test for multiple comparisons (for 1-to-9 months groups), and unpaired t-test for 16 months-old mice. (n=6 mice/group).

**Supplementary Figure 4. Electrocardiographic assessment of *Dsp*$^{S311A}$ mice.**

(**A**) Evaluation of Heart Rate (in beat *per* minute, BPM), in 4 months *Dsp*$^{WT/WT}$, *Dsp*$^{WT/S311A}$ and *Dsp*$^{S311A/S311A}$ male mice. Bars represent s.d. Differences among groups were determined using ordinary one-way ANOVA with uncorrected Fisher's LSD test for multiple comparisons. (n=6 mice/group. **, $p<0.01$). (**B**) ECG-based evaluation of PR interval duration in 4 months *Dsp*$^{WT/WT}$, *Dsp*$^{WT/S311A}$ and *Dsp*$^{S311A/S311A}$ male mice. Bars represent s.d. Differences among groups were determined using ordinary one-way ANOVA with uncorrected Fisher's LSD test for multiple comparisons. (n=6 mice/group. ns, not significant). (**C**) Representative ECG traces from 4-months *Dsp*$^{S311A/S311A}$ and *Dsp*$^{WT/S311A}$ male mice illustrating different types of ectopic beats, including bundle branch block (BBB) (top panel) and couplet (bottom panel). (**D**) Representative ECG traces of 4-months *Dsp*$^{S311A/S311A}$ and *Dsp*$^{WT/S311A}$ male mice showing complex sustained arrhythmias. (**E**) Representative images of ECG traces of 9 months *Dsp*$^{WT/WT}$, *Dsp*$^{WT/S311A}$ and *Dsp*$^{S311A/S311A}$ male mice recorded before (left panel) and after (right panel) noradrenaline administration. Traces show an increased incidence of arrhythmic beats in *Dsp*$^{WT/S311A}$ and *Dsp*$^{S311A/S311A}$ mice upon noradrenaline injection.

**Supplementary Figure 5. Gross heart morphology of *Dsp*$^{S311A}$ mice.**

(**A-D**) Evaluation of different morphometric parameters, including: Right Ventricle (RV, **A**) and Left Ventricle (LV, **B**) wall thickness, and LV Posterior (**C**) and Anterior (**D**) wall thickness. Hearts from *Dsp*$^{WT/WT}$, *Dsp*$^{WT/S311A}$ and *Dsp*$^{S311A/S311A}$ male mice, at different ages, were analyzed. Bars in (**A-D**) represent s.d. Differences among groups were determined using two-way ANOVA with Uncorrected Fisher's LSD test for multiple comparisons. (Each value is the mean of 4 non-consecutive sections from the mid portion of the ventricles. n=3 hearts/group; *, p<0.05; **, p<0.01; ***, p<0.001). (**E-F**) Evaluation of LV (**E**) and RV (**F**) area of $Dsp^{WT/WT}$, $Dsp^{WT/S311A}$ and $Dsp^{S311A/S311A}$ male mice, at different ages. Bars represent s.d. Differences among groups were determined using two-way ANOVA with Uncorrected Fisher's LSD test for multiple comparisons. (Each value is the mean of 4 non-consecutive sections from the mid portion of the ventricles. n=3 hearts/group; *, p<0.05; **, p<0.01; ***, p<0.001). (**G**) Haematoxylin-eosin staining of sections, from the mid portion of the ventricles, from 12 months $Dsp^{WT/WT}$ and $Dsp^{WT/S311A}$ male mice.

**Supplementary Figure 6. Histological analysis of $Dsp^{S311A}$ hearts.**

Left ventricle details of heart sections stained with Haematoxylin-eosin, from the mid portion of the ventricles, of $Dsp^{WT/WT}$, $Dsp^{WT/S311A}$ and $Dsp^{S311A/S311A}$ male mice at different ages.

**Supplementary Figure 7. Cardiomyocyte apoptosis in $Dsp^{S311A}$ hearts.**

(**A**) Representative images of TUNEL assay (green signal) in heart sections from 9 months $Dsp^{WT/WT}$, $Dsp^{WT/S311A}$ and $Dsp^{S311A/S311A}$ male mice. Sections were co-stained with an antibody to $\alpha$-actinin. Nuclei were counterstained with DAPI. Images are details from the LV. (**B-C**) Quantification of the number of TUNEL-positive cardiomyocytes in the right ventricular sub-epicardium of $Dsp^{WT/WT}$, $Dsp^{S311A/S311A}$ (**B**) and $Dsp^{WT/WT}$, $Dsp^{WT/S311A}$ (**C**) male mice, at different ages. Bars represent s.d. Differences among groups were determined using two-way ANOVA with Šídák's test for multiple comparisons. (Each value represents the percentage of TUNEL-positive cardiomyocytes measured in 3 different fields within the sub-epicardial region of the right ventricle, obtained from 4 non-consecutive sections per heart, n=3 hearts/disease stage. **, p<0.01; ****, p<0.0001).

**Supplementary Figure 8. Inflammation in $Dsp^{S311A}$ hearts.**

Confocal immunofluorescence in hearts sections, from the mid portion of the ventricles, of *Dsp*$^{WT/WT}$, *Dsp*$^{WT/S311A}$ and *Dsp*$^{S311A/S311A}$ male mice, at different ages. Sections were stained with an antibody to the macrophage marker, F4/80. Nuclei were counterstained with DAPI.

**Supplementary Figure 9. Fibrotic remodeling of the left ventricle in *Dsp*$^{S311A}$ hearts.**

Heart sections from the mid portion of the ventricles of *Dsp*$^{WT/WT}$, *Dsp*$^{WT/S311A}$ and *Dsp*$^{S311A/S311A}$ male mice, at different ages, co-stained with antibodies to collagen-I and sarcomeric actinin ($\alpha$-actinin). Nuclei were counterstained with DAPI. Images are details of the left ventricle.

**Supplementary Figure 10. Fibrotic remodeling of the interventricular septum in *Dsp*$^{S311A}$ hearts.**

Heart sections from the mid portion of the ventricles of *Dsp*$^{WT/WT}$, *Dsp*$^{WT/S311A}$ and *Dsp*$^{S311A/S311A}$ male mice, at different ages, co-stained with antibodies to collagen-I and sarcomeric actinin ($\alpha$-actinin). Nuclei were counterstained with DAPI. Images are details of the interventricular septum.

**Supplementary Figure 11. Fibrotic remodeling of the right ventricle in *Dsp*$^{S311A}$ hearts.**

Heart sections from the mid portion of the ventricles of *Dsp*$^{WT/WT}$, *Dsp*$^{WT/S311A}$ and *Dsp*$^{S311A/S311A}$ male mice, at different ages, were co-stained with antibodies to collagen-I and sarcomeric actinin ($\alpha$-actinin). Nuclei were counterstained with DAPI. Images are details of the right ventricle.

**Supplementary Figure 12. Quantification of collagen-I deposition in *Dsp*$^{S311A}$ hearts.**

(**A-B**) Quantification of the percentage of myocardial area of the right ventricle (RV) (**A-B**) and interventricular septum (IVS) (**C-D**) occupied by collagen-I, in hearts from *Dsp*$^{WT/WT}$ (**A-** **-D**), *Dsp*$^{S311A/S311A}$ (**A, C**) and *Dsp*$^{WT/S311A}$ (**B, D**) male mice, at different ages. Differences among groups were determined using two-way ANOVA with uncorrected Fisher's LSD test for multiple comparisons. (Each value represents the mean of 4 non-consecutive sections from 3 different hearts. n=3 mice/group. **, $p<0.01$; ***, $p<0.001$; ****, $p<0.0001$).

**Supplementary Figure 13. Tissue calcification in *Dsp*$^{S311A}$ hearts.**

(**A-D**) Von Kossa staining in heart sections from the mid portion of the ventricles of 9 months *Dsp*$^{WT/S311A}$ (**A, C**) and *Dsp*$^{S311A/S311A}$ (**B, D**) male mice. Images (**C-D**) are high magnifications of the black boxes in (**A-B**) respectively.

**Supplementary Figure 14. Ultrastructural assessment of *Dsp*$^{S311A}$ hearts.**

(**A**) Transmission Electron Microscopy (TEM) of thin heart sections from 1-month *Dsp*$^{WT/WT}$, *Dsp*$^{WT/S311A}$ and *Dsp*$^{S311A/S311A}$ male mice. Desmosomes (DS, arrows); Fasciae adherens (FA, arrowheads). (**B**) Evaluation of mean FA intercellular cleft width in 1 and 9 months *Dsp*$^{WT/WT}$, *Dsp*$^{WT/S311A}$ and *Dsp*$^{S311A/S311A}$ male mice. Bars represent s.d. Differences among groups were determined using two-way ANOVA with Tukey's test for multiple comparisons. (From 4 to 40 fields from 4 different hearts/group; ****, $p<0.0001$).

**Supplementary Figure 15. Connexin-43 localization in the left ventricle of *Dsp*$^{S311A}$ hearts.**

Confocal immunofluorescence of heart ventricular sections from *Dsp*$^{WT/WT}$, *Dsp*$^{WT/S311A}$ and *Dsp*$^{S311A/S311A}$ male mice, at different ages, co-stained with antibodies to connexin-43 (cx-43) and sarcomeric actinin ($\alpha$-actinin). Nuclei were counterstained with DAPI. Images are details of the left ventricle.

**Supplementary Figure 16. Connexin-43 localization in the interventricular septum of *Dsp*$^{S311A}$ hearts.**

Confocal immunofluorescence of heart ventricular sections from *Dsp*$^{WT/WT}$, *Dsp*$^{WT/S311A}$ and *Dsp*$^{S311A/S311A}$ male mice, at different ages, co-stained with antibodies to connexin-43 (cx-43), sarcomeric actinin ($\alpha$-actinin). Nuclei were counterstained with DAPI. Images are details of the interventricular septum.

**Supplementary Figure 17. Connexin-43 localization in the right ventricle of *Dsp*$^{S311A}$ hearts.**

Confocal immunofluorescence of heart ventricular sections from *Dsp*$^{WT/WT}$, *Dsp*$^{WT/S311A}$ and *Dsp*$^{S311A/S311A}$ male mice, at different ages, co-stained with antibodies to connexin-43 (cx-43), sarcomeric actinin ($\alpha$-actinin). Nuclei were counterstained with DAPI. Images are details of the right ventricle.

**Supplementary Figure 18. β-catenin signaling in *Dsp*$^{S311A}$ hearts.**

(**A-B**) Confocal immunofluorescence of heart ventricular sections from 4 months *Dsp*$^{WT/WT}$ (**A**) and *Dsp*$^{S311A/S311A}$ (**B**) male mice, stained with an antibody to the β-catenin interactor, LEF1. Nuclei were counterstained with DAPI. (**C-D**) Confocal immunofluorescence of heart ventricular sections from 9 (**C**) and 16 (**D**) months *Dsp*$^{WT/S311A}$ male mice co-stained with antibodies to β-catenin and $\alpha$-actinin. Nuclei were counterstained with DAPI.

**Supplementary Figure 19. Effect of exercise on arrhythmia and heart remodeling in *Dsp*$^{S311A}$ hearts.**

(**A**) Representative ECG traces from 4 months sedentary *vs.* exercised *Dsp*$^{WT/S311A}$ and *Dsp*$^{S311A/S311A}$ male mice, showing complex sustained arrhythmias upon exercise. (**B-C**) Representative images of TUNEL assay (green signal) in heart sections from 4 months sedentary (sed.) *vs.* exercised (exe.) *Dsp*$^{WT/S311A}$ (**B**) and *Dsp*$^{S311A/S311A}$ (**C**) male mice. Sections were co-stained with an antibody to $\alpha$-actinin. Nuclei were counterstained with DAPI. Images are details from the LV. (**D-E**) Haematoxylin-eosin staining of heart sections, from the mid portion of the ventricles, of 4 months sedentary (**E**) *vs.* exercised (**F**) *Dsp*$^{S311A/S311A}$ male mice. Right images in (**D-E**) are high magnifications of the black boxes in composite images.

| Name of the gene | Direction | Sequence |
|---|---|---|
| *Dsp* | Forward | CAAAAGCAGGCTTTAGAGGCAT |
| | Reverse | CTTCACCAGCAGGCTCTCTC |
| *Dsp* | Forward | GCCGCTCTCAACTATGTCTG |
| | Reverse | AGAACACTGACTGCTCTTCC |
| *Dsg2 C-term* | Forward | GTGCGCCATTCAGCTTCTCC |
| | Reverse | AGTGAGCTGAAGGACCTGCC |
| *Dsg2 N-term* | Forward | TGCTTGGACTTTGGAAACGGAC |
| | Reverse | TTCTGGACAGGTCTTCGCCC |
| *Jup* | Forward | ATCCACGCCATCCTGAGAGC |
| | Reverse | CACAGAGCCAGGTTCCGGAT |
| *Dsc2* | Forward | CTGTGGGATCTATGCGCTCC |
| | Reverse | TCCATCAGGTTCACTCTGCC |
| *Pkp2* | Forward | CCACAGCCTCTGCTTGCTAT |
| | Reverse | CTTGTTGGGGGCATAGCCTT |
| *Cx43* | Forward | AGTGAAAGAGAGGTGCCCAGA |
| | Reverse | GTGGAGTAGGCTTGGACCTT |
| *Gapdh* | Forward | CACCATCTTCCAGGAGCGAG |
| | Reverse | CCTTCTCCATGGTGGTGAAGAC |

**Supplementary Table 1.** Oligos used in this study

| Target | Supplier | Host | Catalog Number | Dilution | Method |
|---|---|---|---|---|---|
| α-actinin | Sigma-Aldrich | Mouse | A7732 | 1:200 | IF |
| β-actin | Sigma-Aldrich | Rabbit | A5060 | 1:2500 | WB |
| β-catenin | Abcam | Rabbit | AB16051 | 1:100 | IF |
| CD4 | BD Pharmingen | Rat | 553730 | 1:50 | FC |
| CD8a | BioLegend | Rat | 100705 | 1:50/1:50 | IF/FC |
| Collagen I (Col1a1) | OriGene | Rabbit | BP8003S | 1:80 | IF |
| Collagen I (Col1a1) | Abcam | Mouse | AB6308 | 1:100 | IF |
| Connexin-43 | Merck Millipore | Rabbit | AB1727 | 1:100 | IF |
| Desmocollin-2 | Santa Cruz Biotechnology | Mouse | SC-70994 | 1:50 | IF |
| Desmoglein-2 | Abcam | Rabbit | AB150372 | 1:1000 | WB |
| Desmoglein-2 | Abcam | Rabbit | AB85632 | 1:50 | IF |
| Desmoplakin | Progen | Mouse | 61003 | 1:50/1:200 | IF/WB |
| F4/80 | Abcam | Rat | AB6640 | 1:150 | IF |
| Gamma Catenin (Plakoglobin (JUP)) | Abcam | Rabbit | AB15153 | 1:100/1:500 | IF/WB |
| GAPDH | Cell Signaling Technology | Mouse | 97166 | 1:10000 | WB |
| LEF-1 | Abcam | Rabbit | AB137872 | 1:100 | IF |
| Perilipin-1 | Cell Signaling Technology | Rabbit | 9349 | 1:200 | IF |

| | | | | | |
|---|---|---|---|---|---|
| Plakophilin-2 | Progen | Mouse | 651101 | 1:50/1:1000 | IF/WB |

**Supplementary Table 2.** Primary antibodies used in this study.

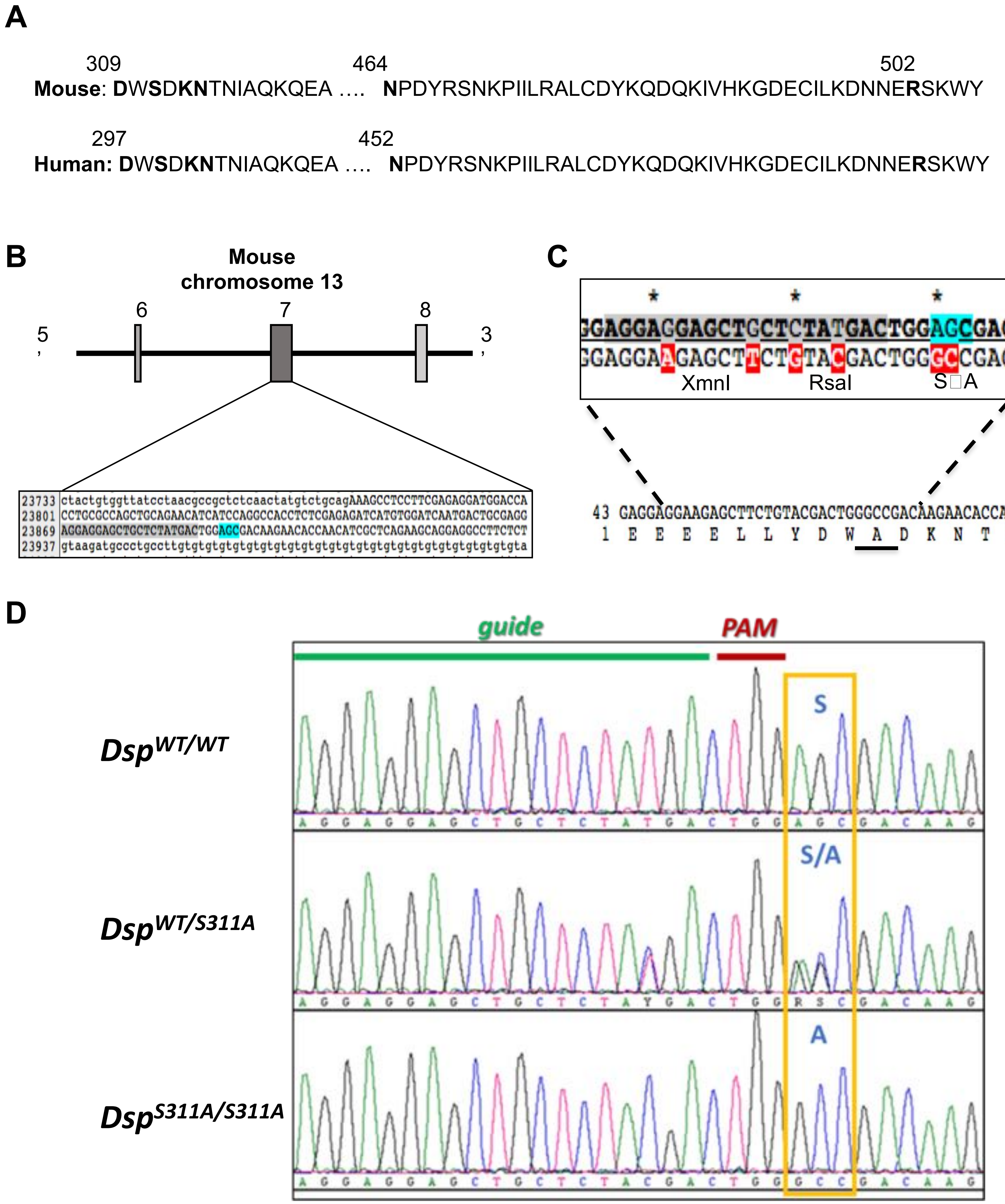

**Supplementary Figure 1**

A

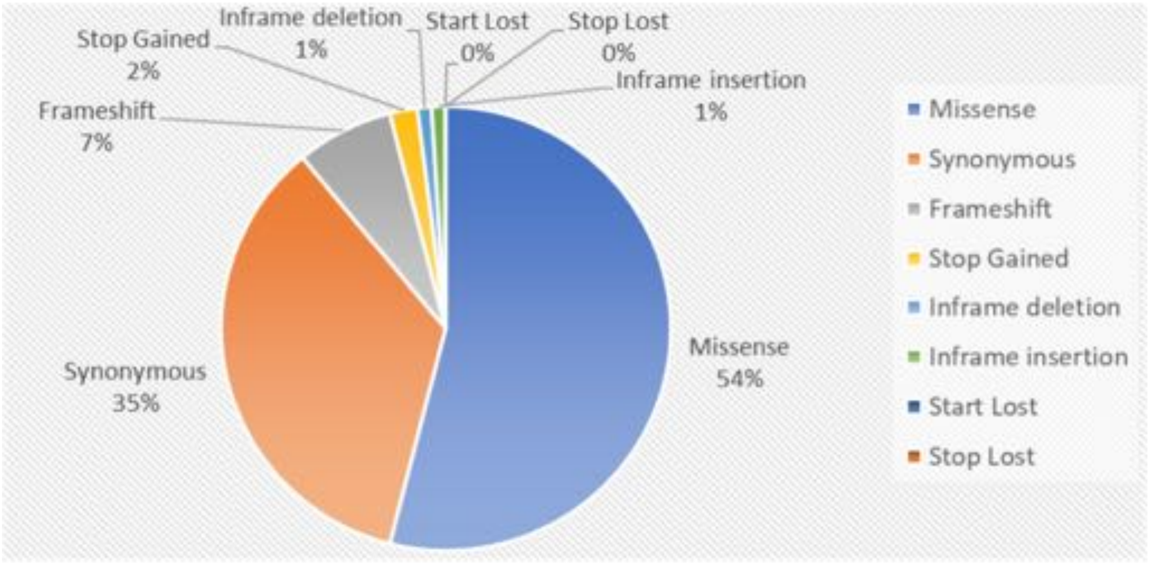

B

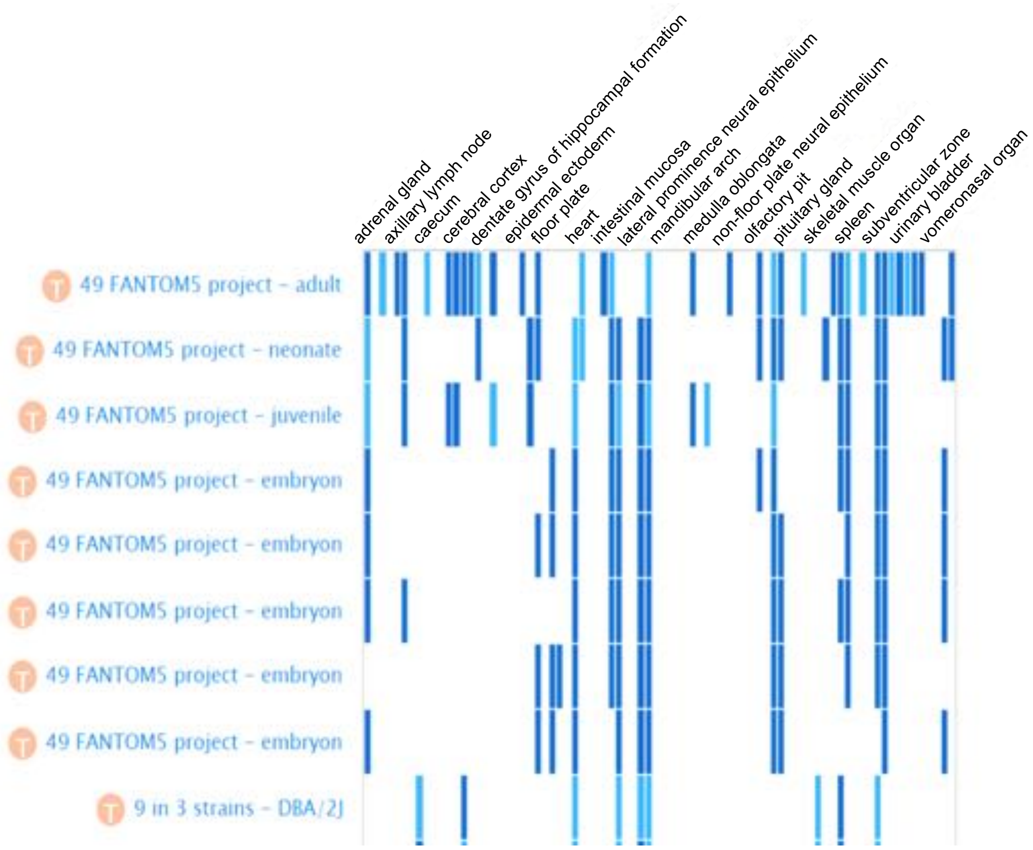

C

| Gene Symbol | Gene Name |
|---|---|
| Hjurp | Holliday Junction Recognition Protein |
| Zfp984 | Zinc Finger Protein 984 |
| LOC108168049 | // |
| Muc4 | Mucin 4, Cell Surface Associated |
| Gm8369 | |
| Gm15266 | |
| Aspm | Abnormal Spindle Microtubule Assembly |
| Sptbn5 | Spectrin Beta, Non-Erythrocytic 5 |
| Zfp513 | Zinc Finger Protein 513 |
| Krtap4-16 | Keratin Associated Protein 4-16 |
| LOC108168049 | // |

**Supplementary Figure 2**

**A**

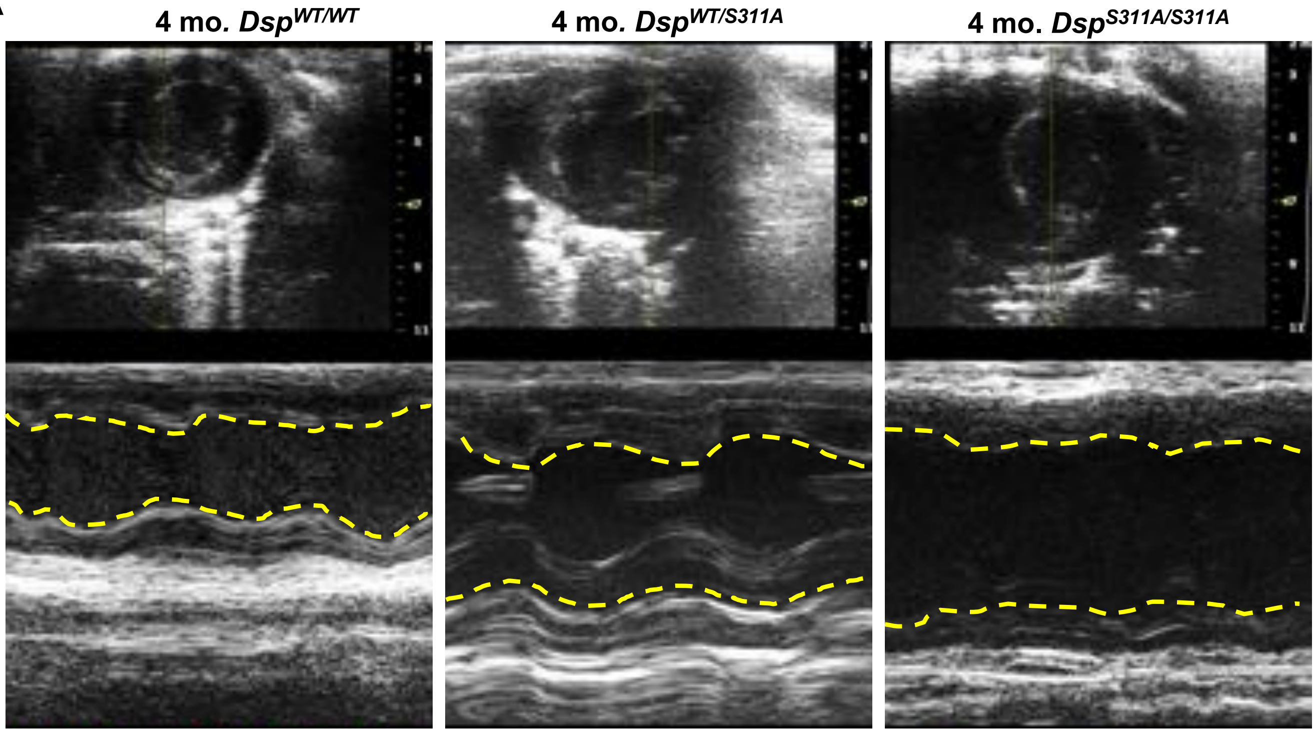

**B**

| Genotype | Age (months) | EF (%) | FS (%) |
|---|---|---|---|
| *Dsp*$^{WT/WT}$ | 1 | $58.50 \pm 2.15$ | $29.64 \pm 1.21$ |
| *Dsp*$^{S311A/S311A}$ | 1 | $69.41 \pm 7.44^{A,B}$ | $36.57 \pm 5.01^{A,B}$ |
| *Dsp*$^{WT/S311A}$ | 1 | $70.74 \pm 8.64^{A}$ | $41.57 \pm 7.88^{A,*}$ |
| *Dsp*$^{WT/WT}$ | 2 | $68.51 \pm 3.67$ | $34.14 \pm 5.97$ |
| *Dsp*$^{S311A/S311A}$ | 2 | $54.13 \pm 4.79^{A,B,*}$ | $27.69 \pm 3.02^{A,B,\S}$ |
| *Dsp*$^{WT/S311A}$ | 2 | $63.50 \pm 5.99^{A}$ | $41.57 \pm 7.88^{A}$ |
| *Dsp*$^{WT/WT}$ | 4 | $62.96 \pm 7.30$ | $31.02 \pm 5.47$ |
| *Dsp*$^{S311A/S311A}$ | 4 | $42.32 \pm 15.76^{A,B,*,\S}$ | $21.19 \pm 9.52^{A,B,\S}$ |
| *Dsp*$^{WT/S311A}$ | 4 | $62.01 \pm 15.14^{A}$ | $33.84 \pm 11.40^{A}$ |
| *Dsp*$^{WT/WT}$ | 6 | $61.54 \pm 6.35$ | $32.66 \pm 4.27$ |
| *Dsp*$^{S311A/S311A}$ | 6 | $33.48 \pm 11.39^{A,B,*,\S}$ | $15.99 \pm 6.15^{A,B,*,\S}$ |
| *Dsp*$^{WT/S311A}$ | 6 | $59.69 \pm 15.86^{A}$ | $34.87 \pm 12.93^{A}$ |
| *Dsp*$^{WT/WT}$ | 9 | $61.45 \pm 5.03$ | $31.63 \pm 4.44$ |
| *Dsp*$^{S311A/S311A}$ | 9 | $26.77 \pm 9.99^{A,B,*,\S}$ | $12.25 \pm 4.74^{A,B,*,\S}$ |
| *Dsp*$^{WT/S311A}$ | 9 | $58.04 \pm 11.31^{A}$ | $30.59 \pm 6.91^{A}$ |
| *Dsp*$^{WT/WT}$ | 16 | $62.45 \pm 4.67$ | $19.08 \pm 10.68$ |
| *Dsp*$^{WT/S311A}$ | 16 | $48.78 \pm 2.35^{A,*}$ | $11.76 \pm 4.61^{A}$ |

$^{A}$ All groups compared with *Dsp*$^{WT/WT}$, $^{B}$ All groups compared with *Dsp*$^{WT/S311A}$, $^{*}$ All groups significantly different when compared with *Dsp*$^{WT/WT}$, $^{\S}$ All groups significantly different when compared with *Dsp*$^{WT/S311A}$

**Supplementary Figure 3**

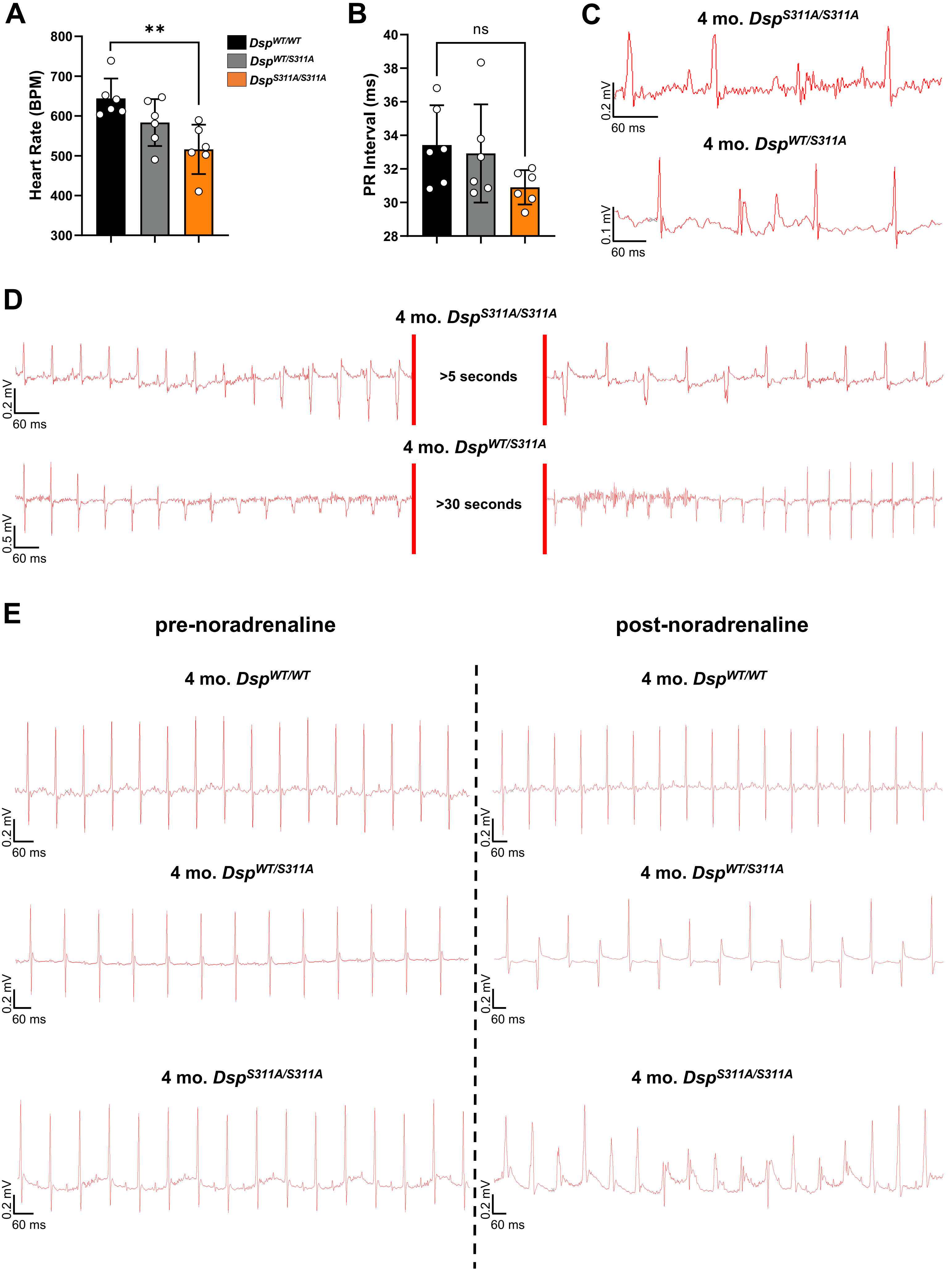

**Supplementary Figure 4**

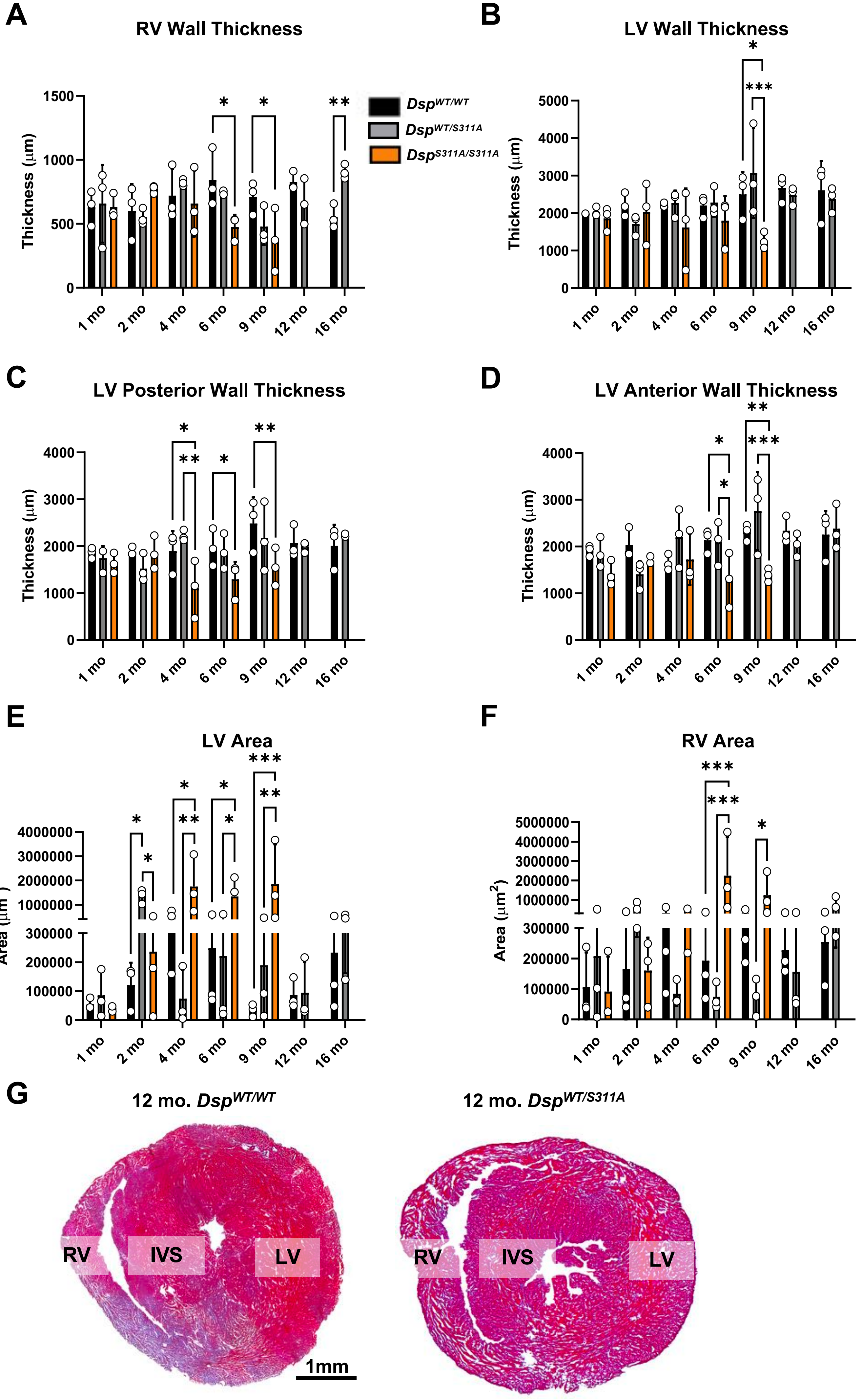

Supplementary Figure 5

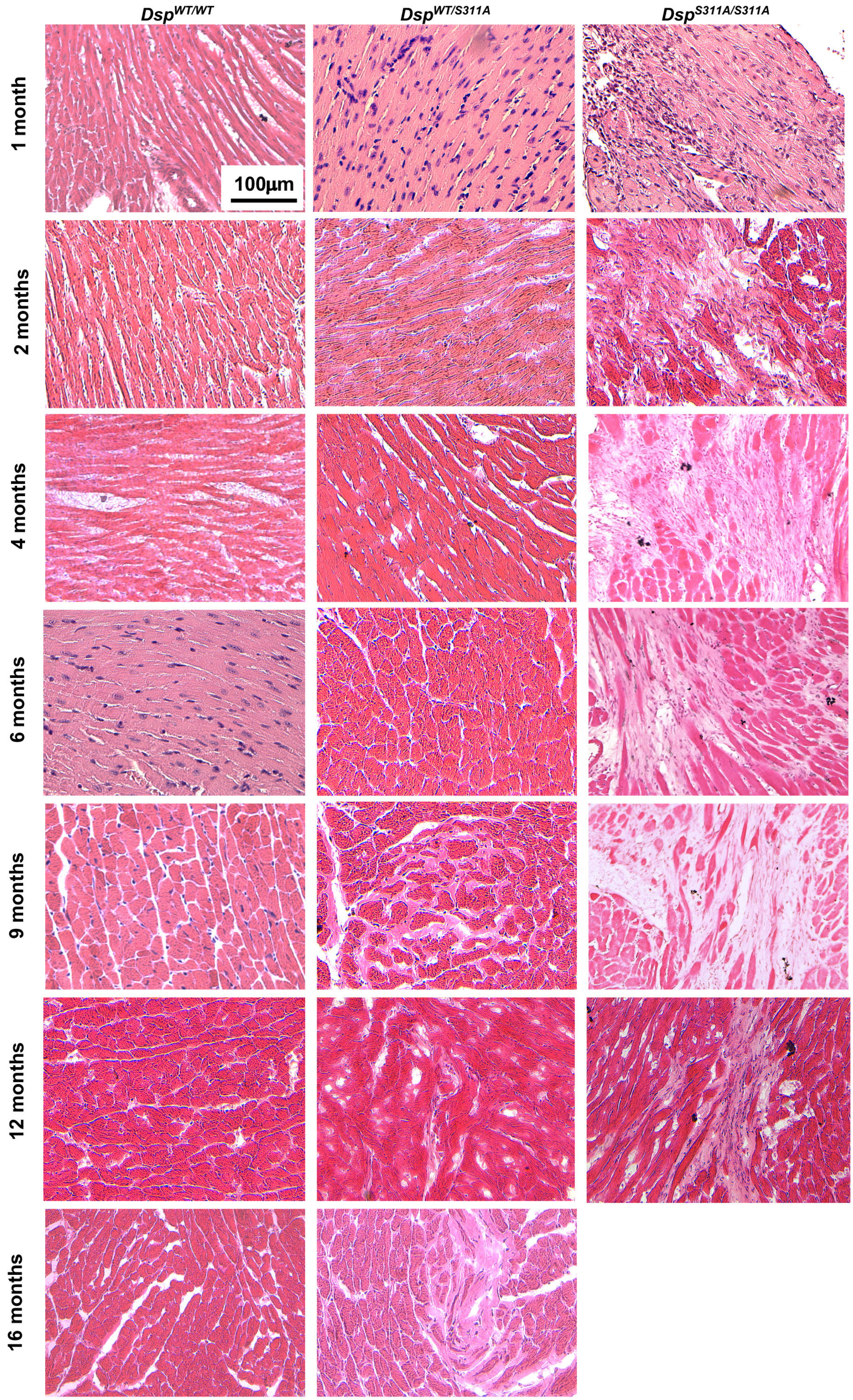

**Supplementary Figure 6**

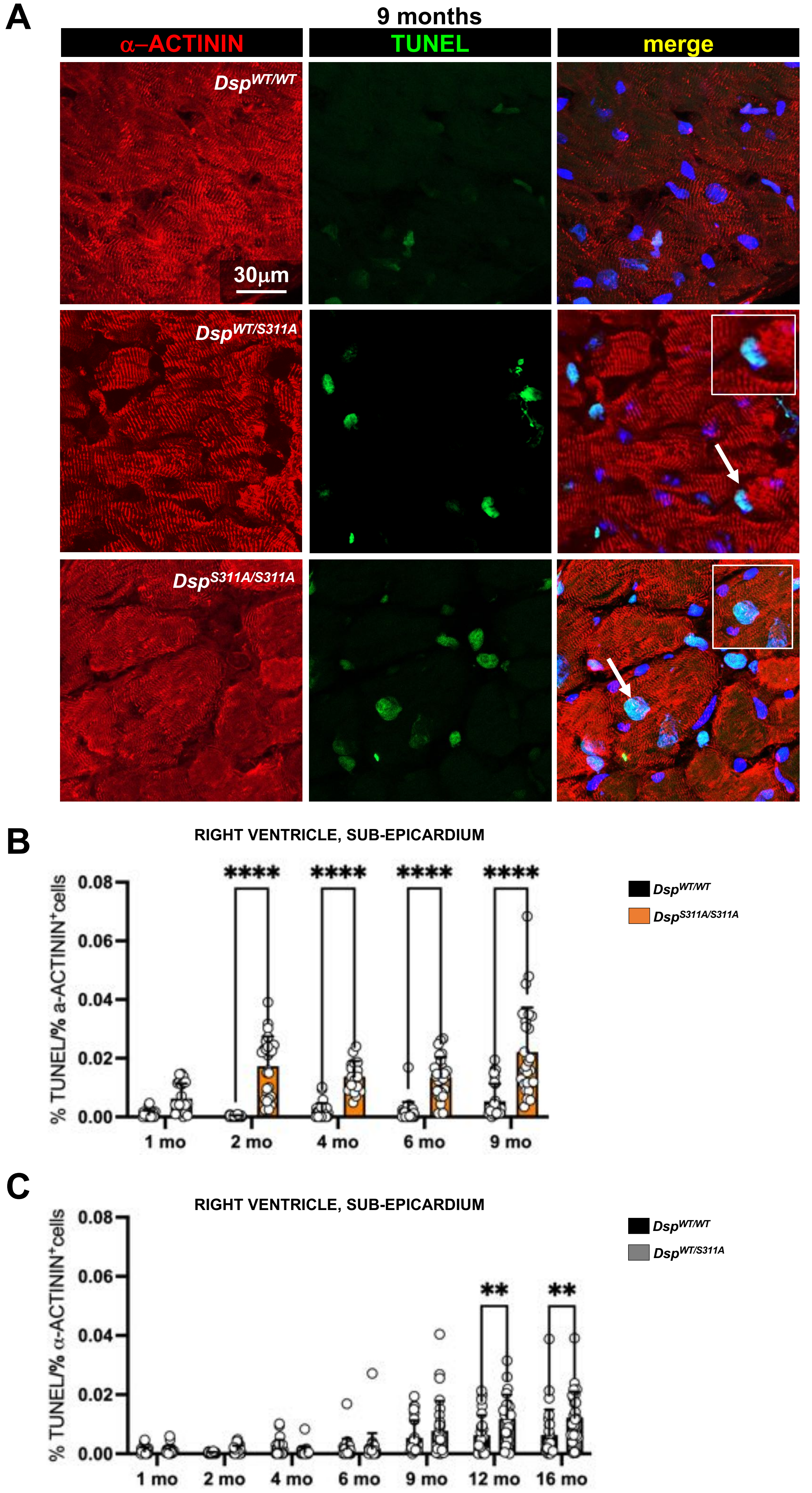

### Supplementary Figure 7

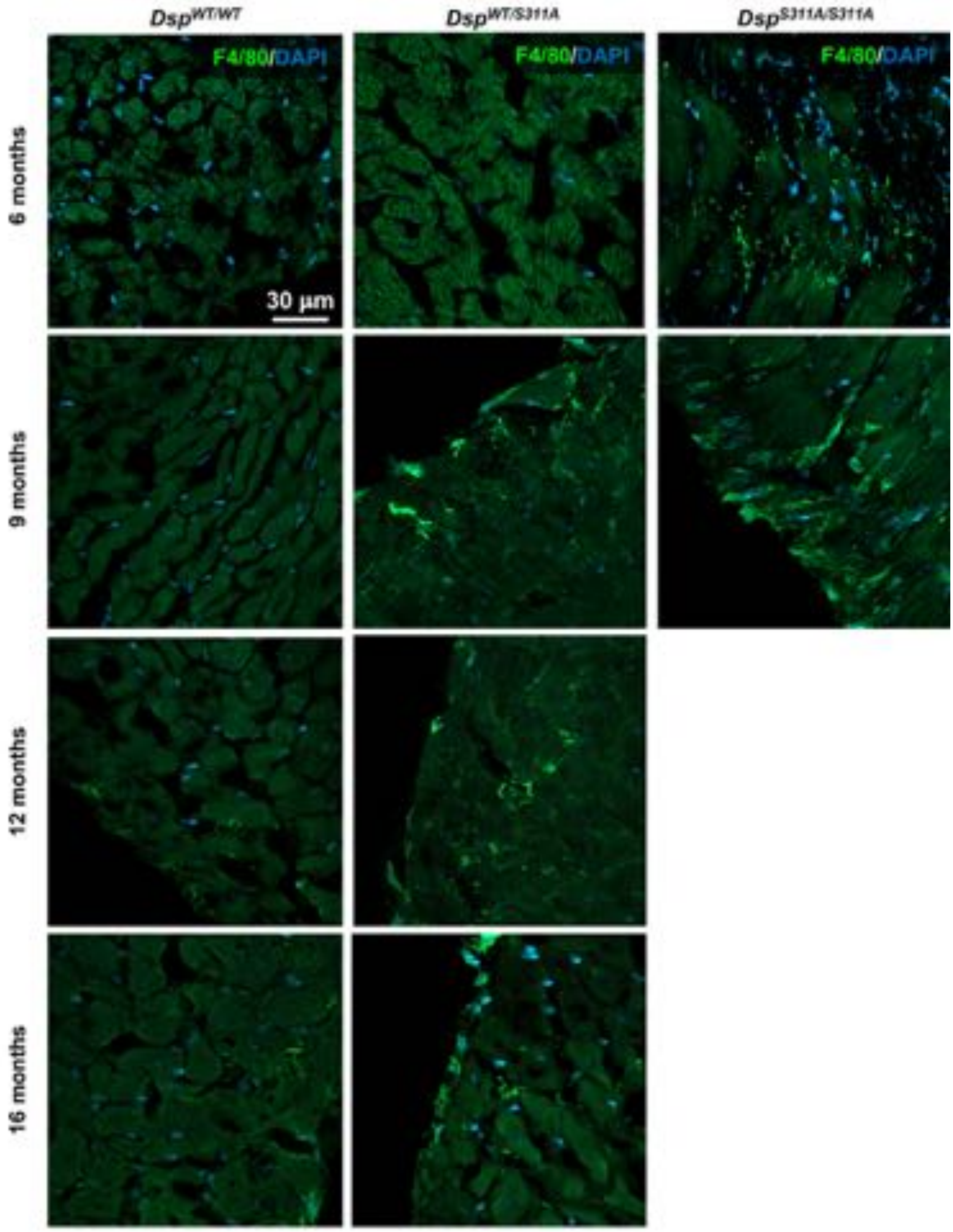

**Supplementary Figure 8**

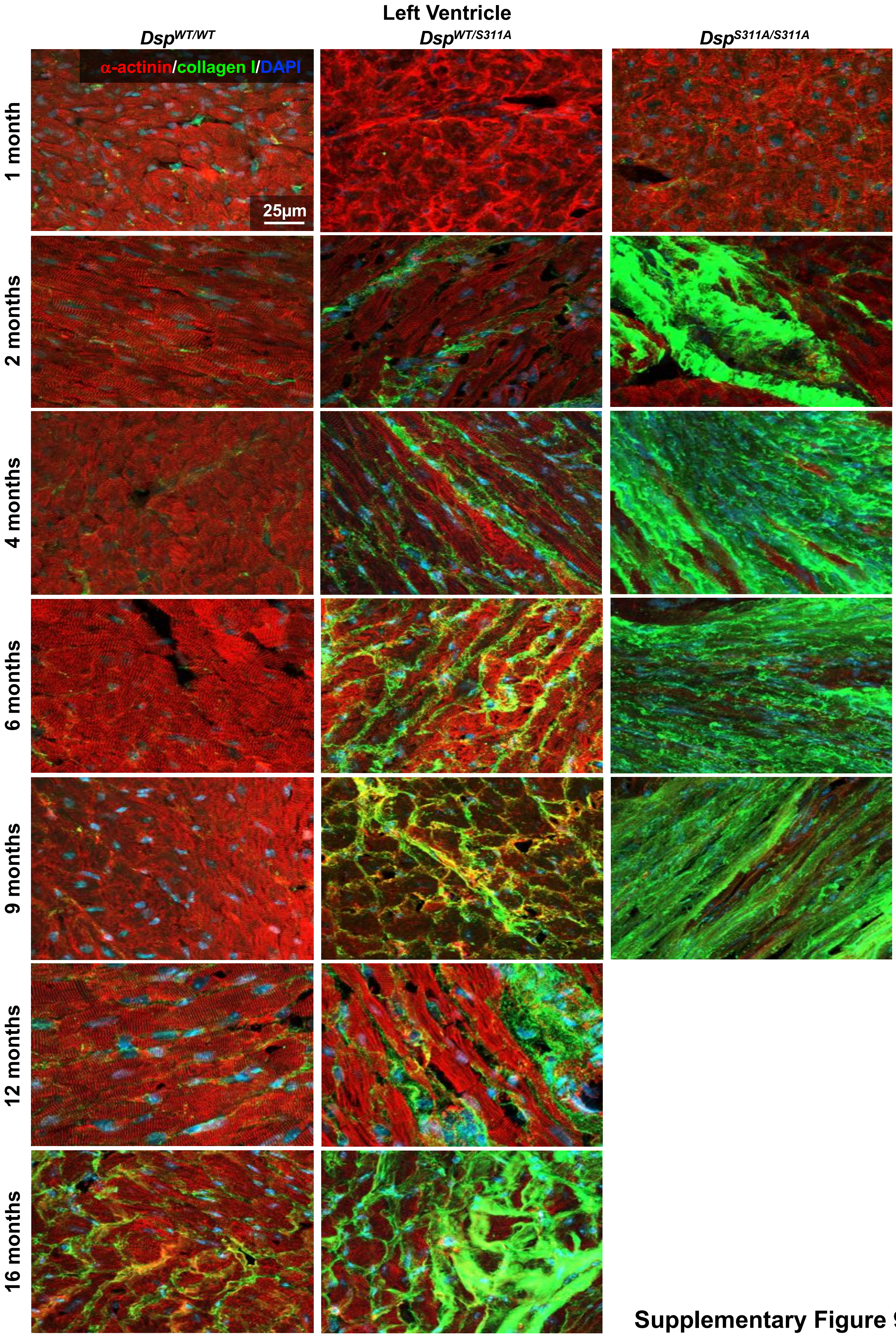

**Supplementary Figure 9**

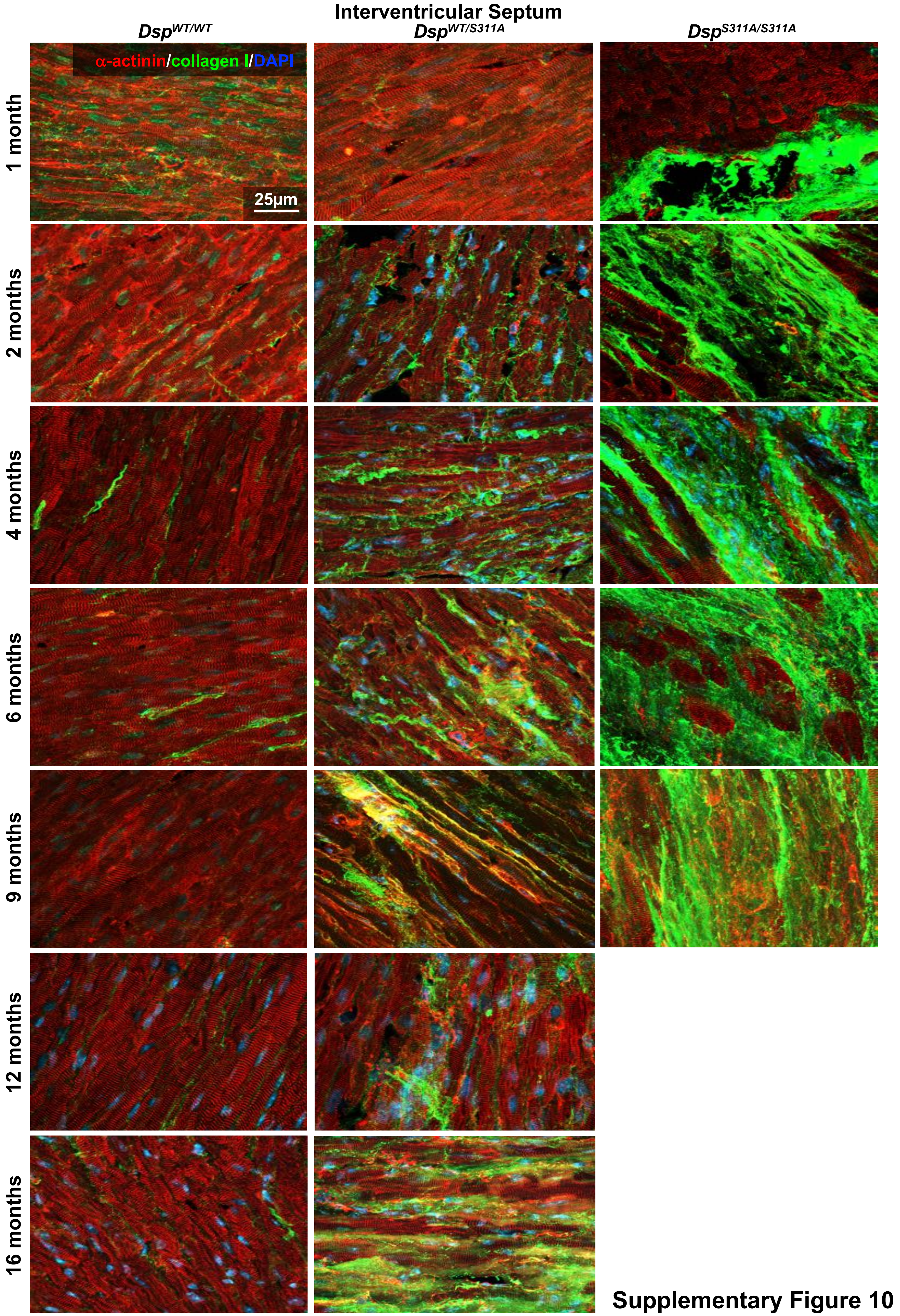

Supplementary Figure 10

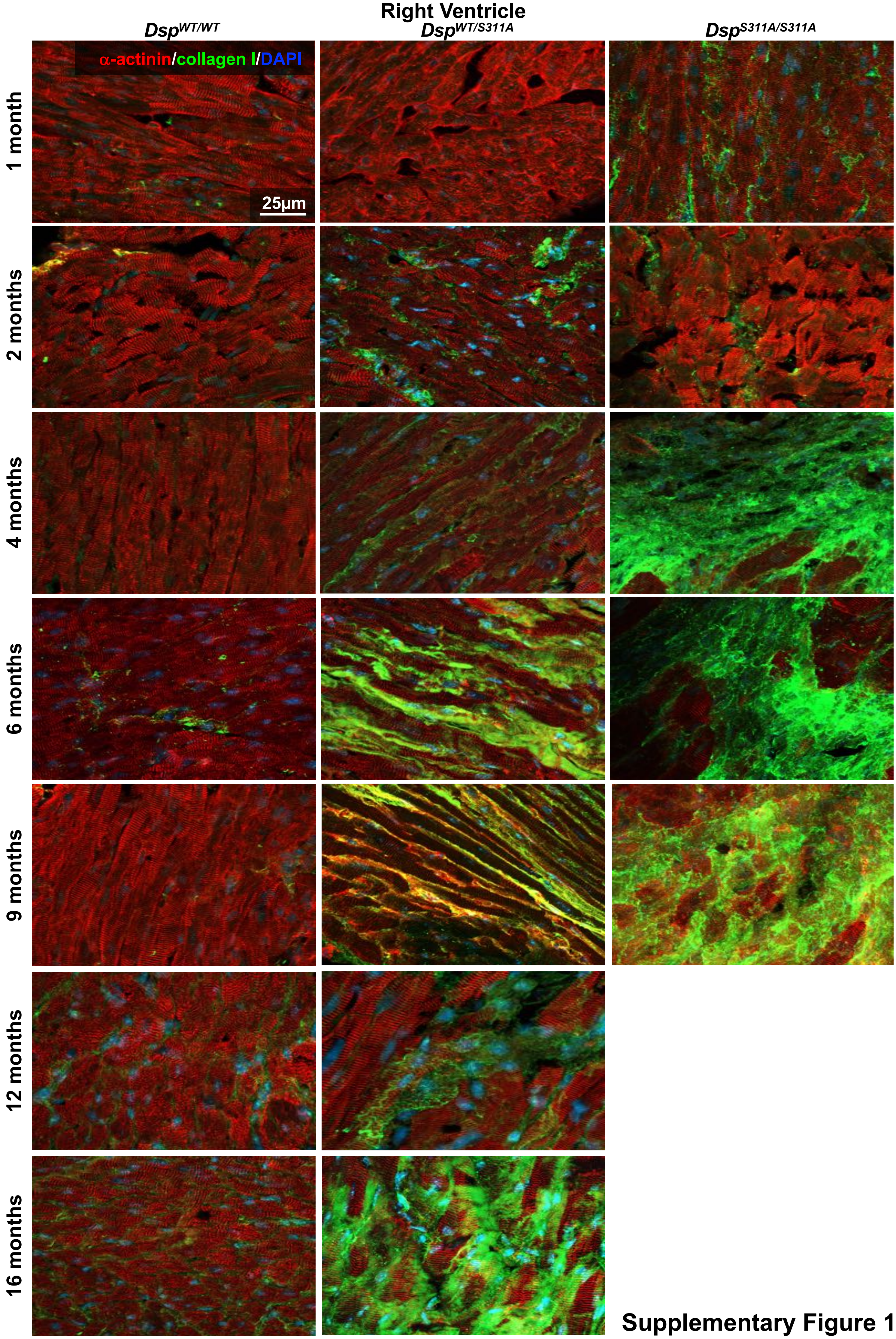

Supplementary Figure 11

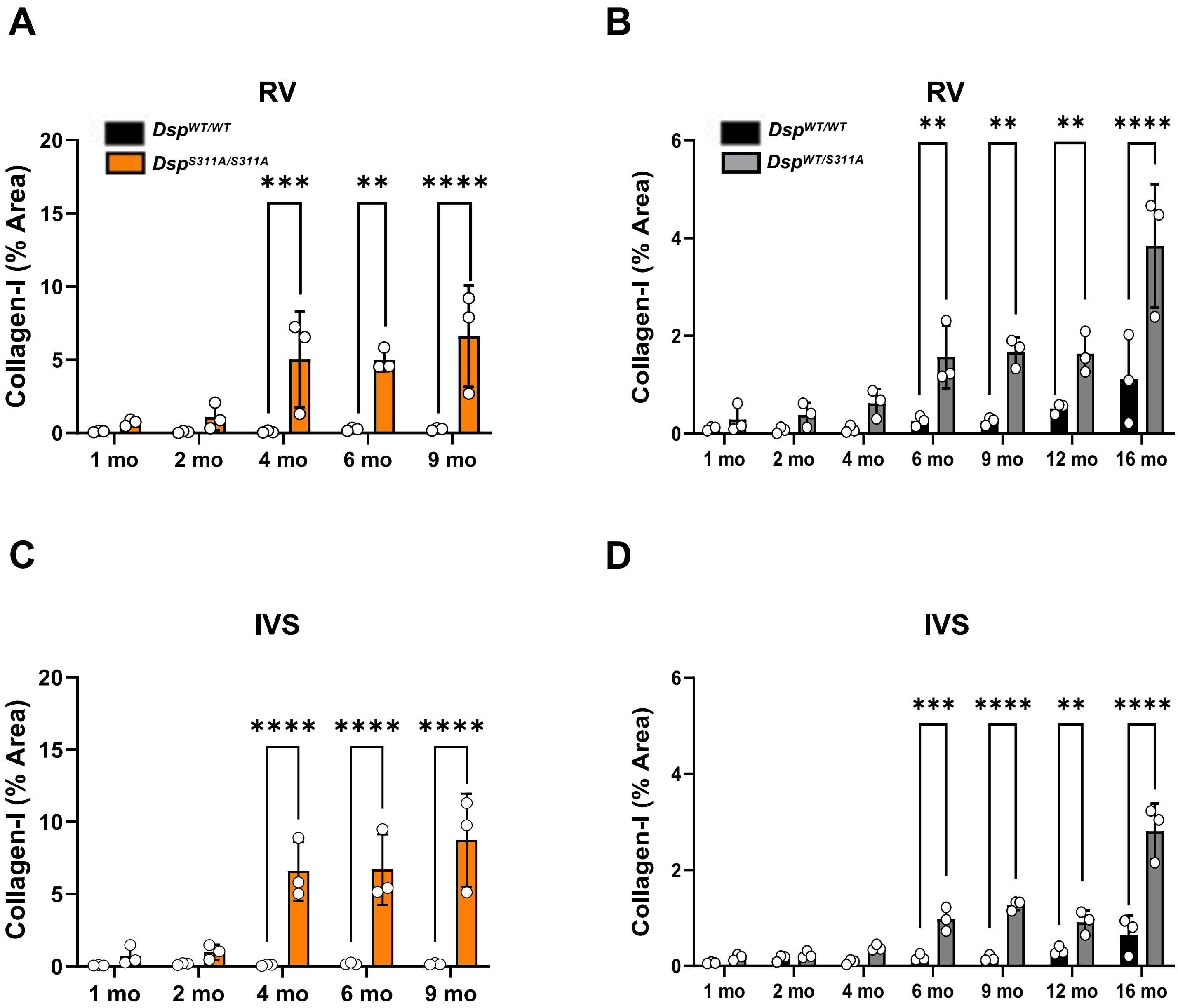

**Supplementary Figure 12**

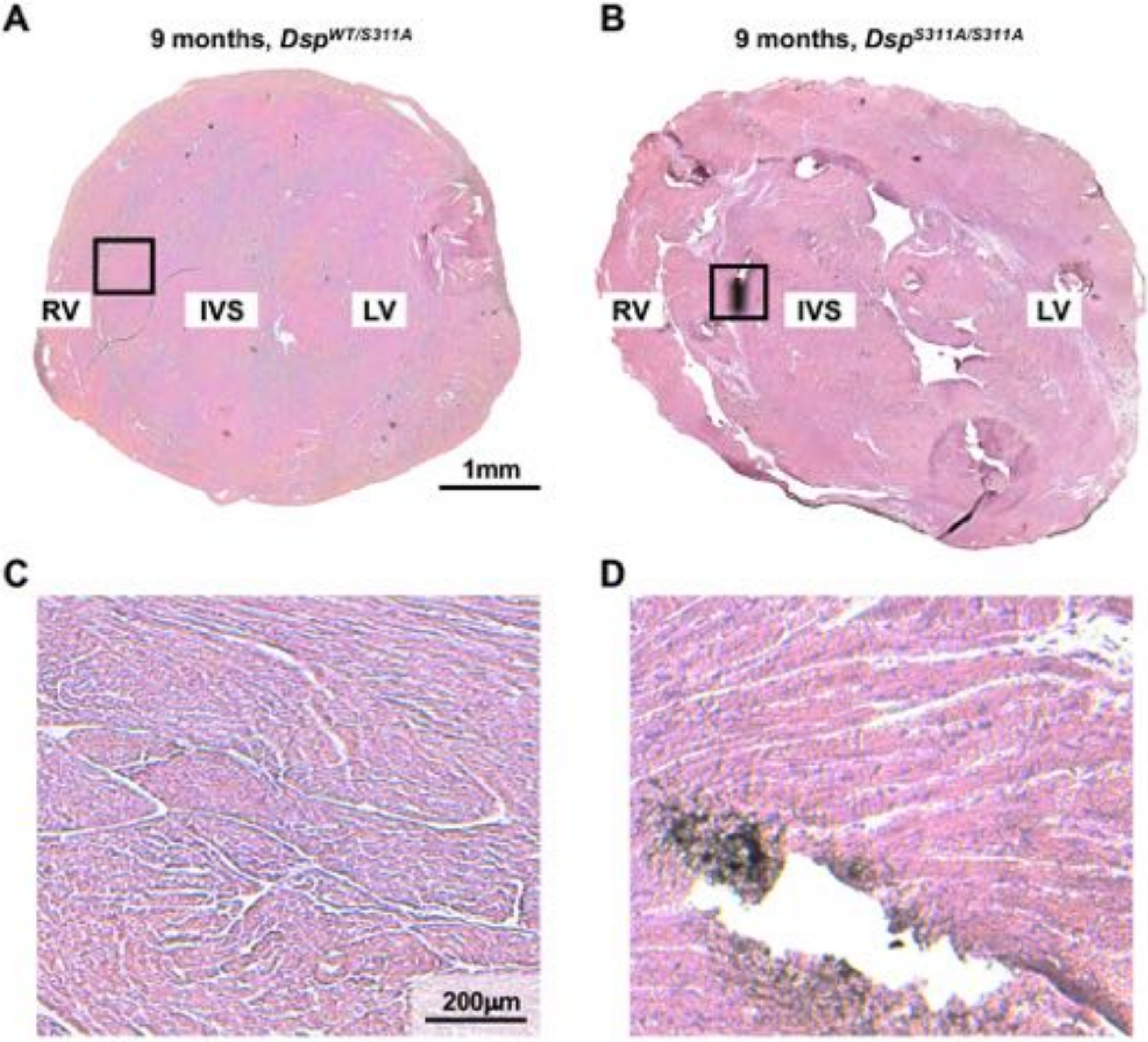

**Supplementary Figure 13**

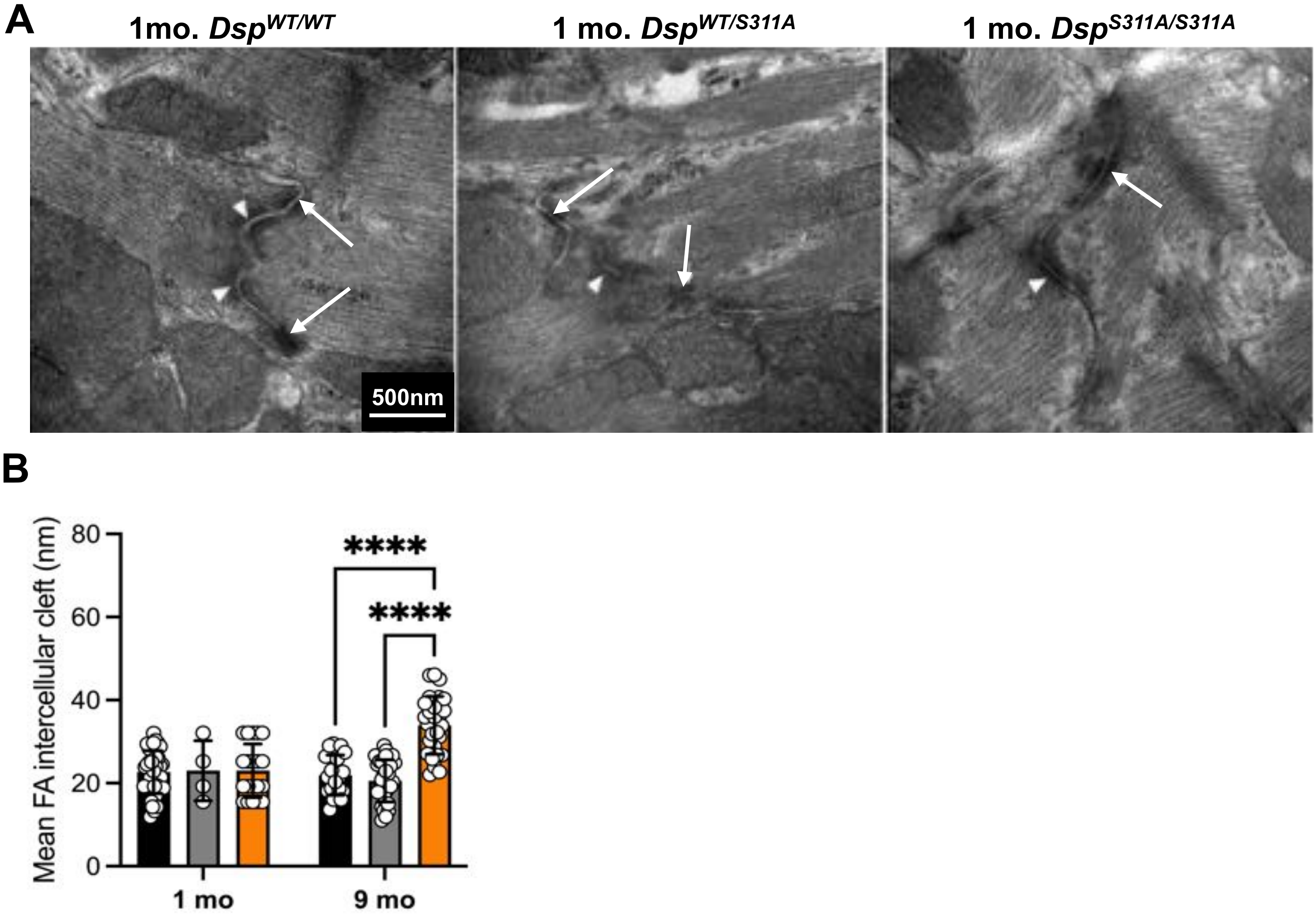

**Supplementary Figure 14**

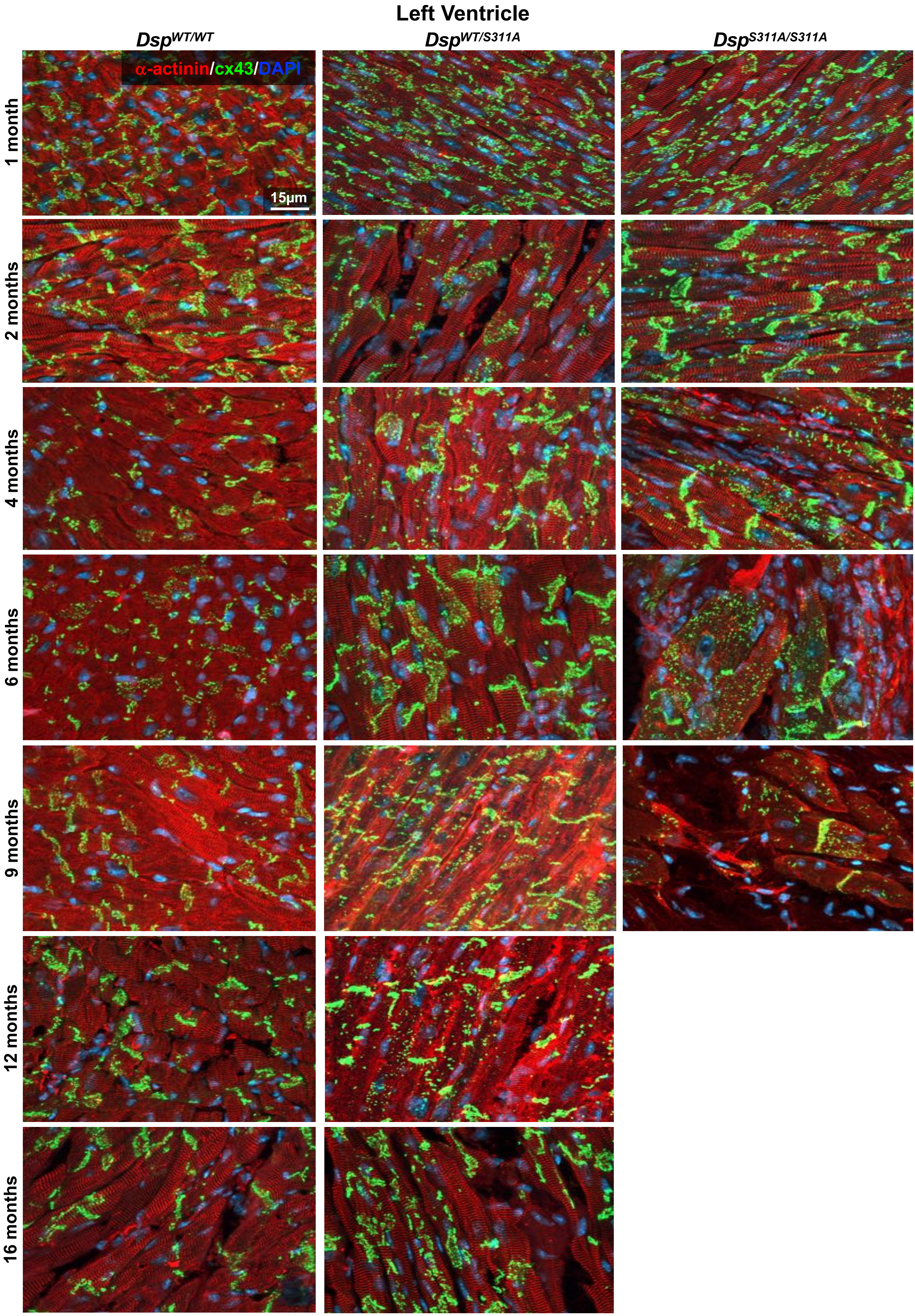

### Supplementary Figure 15

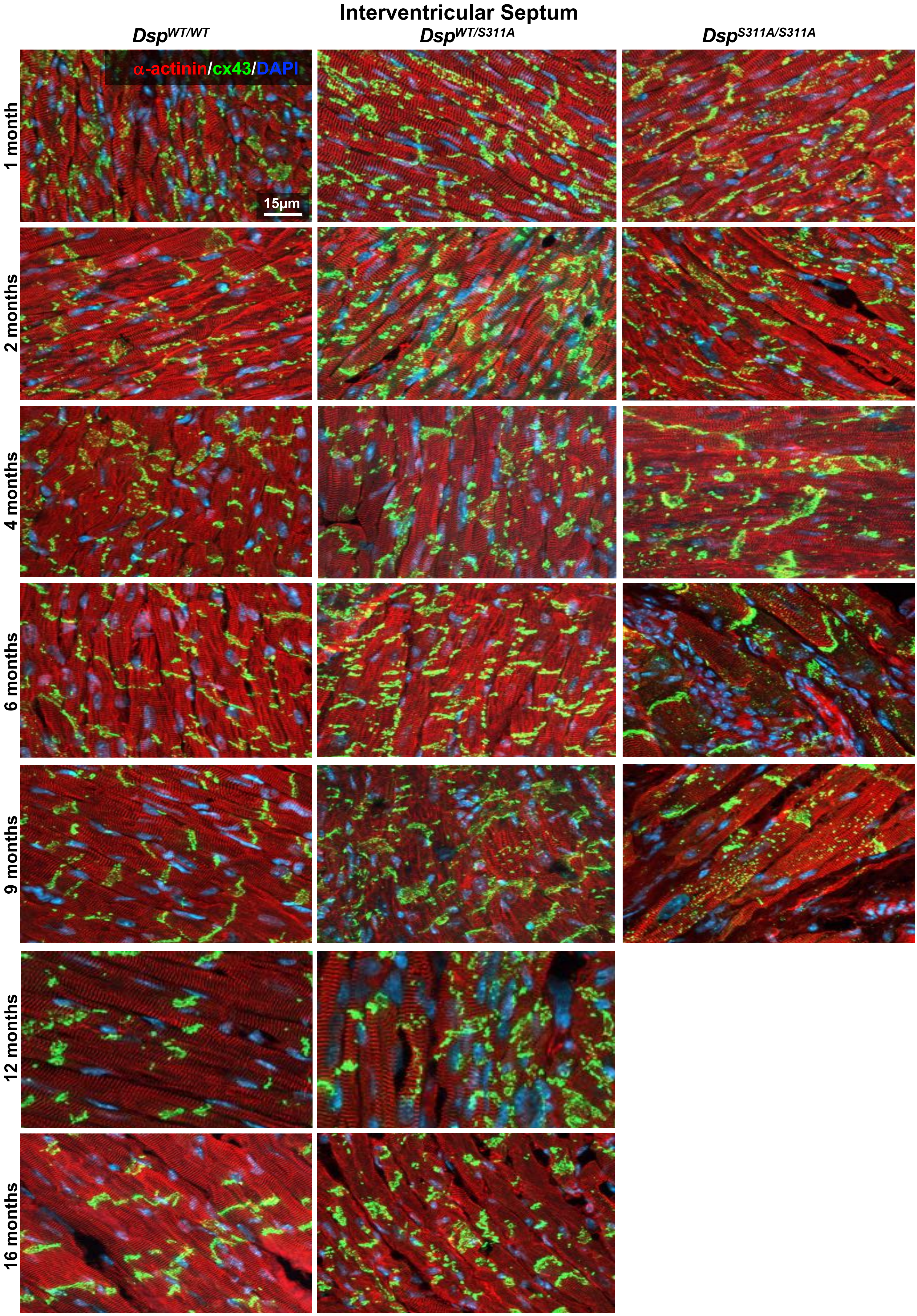

### Supplementary Figure 16

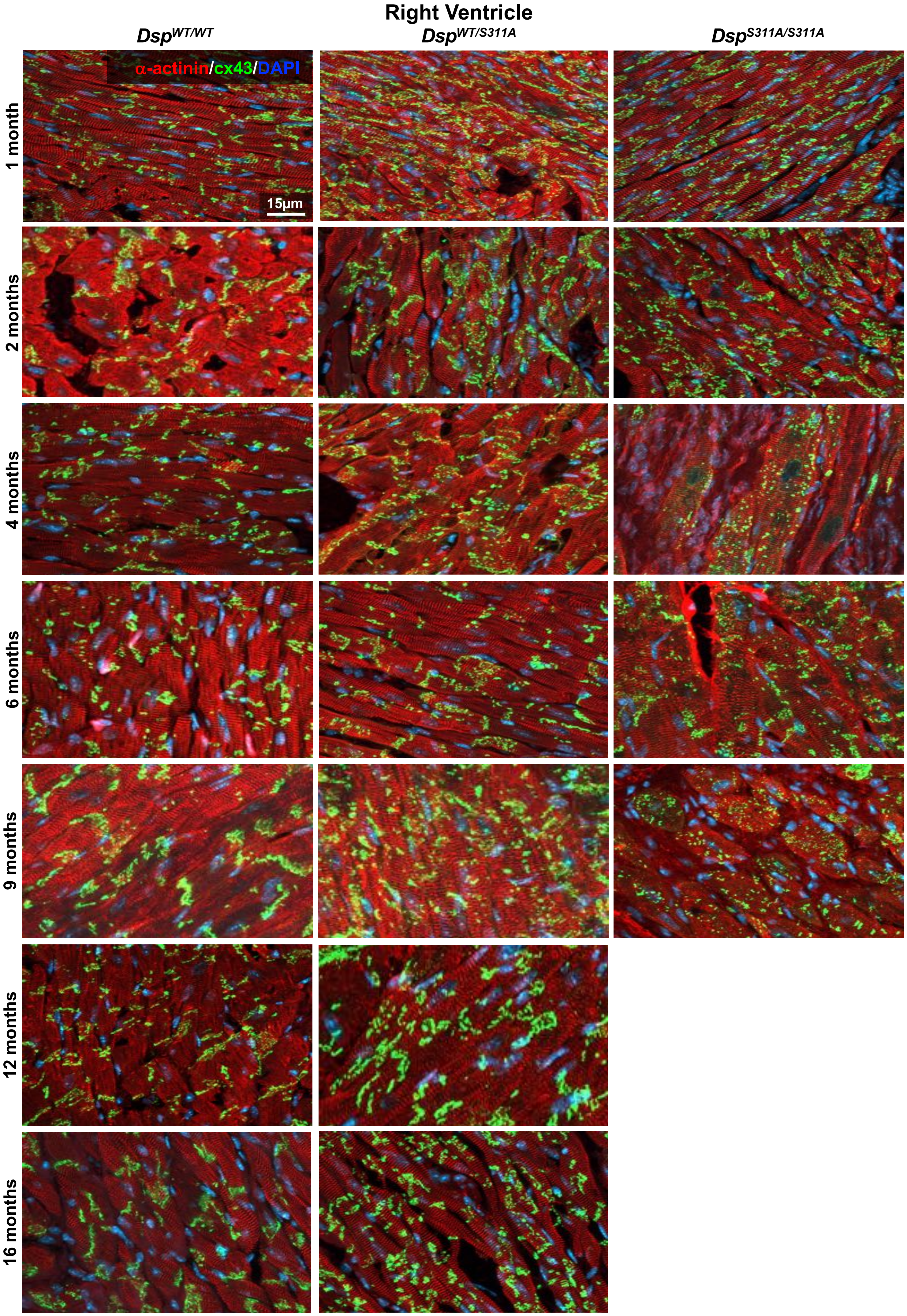

**Supplementary Figure 17**

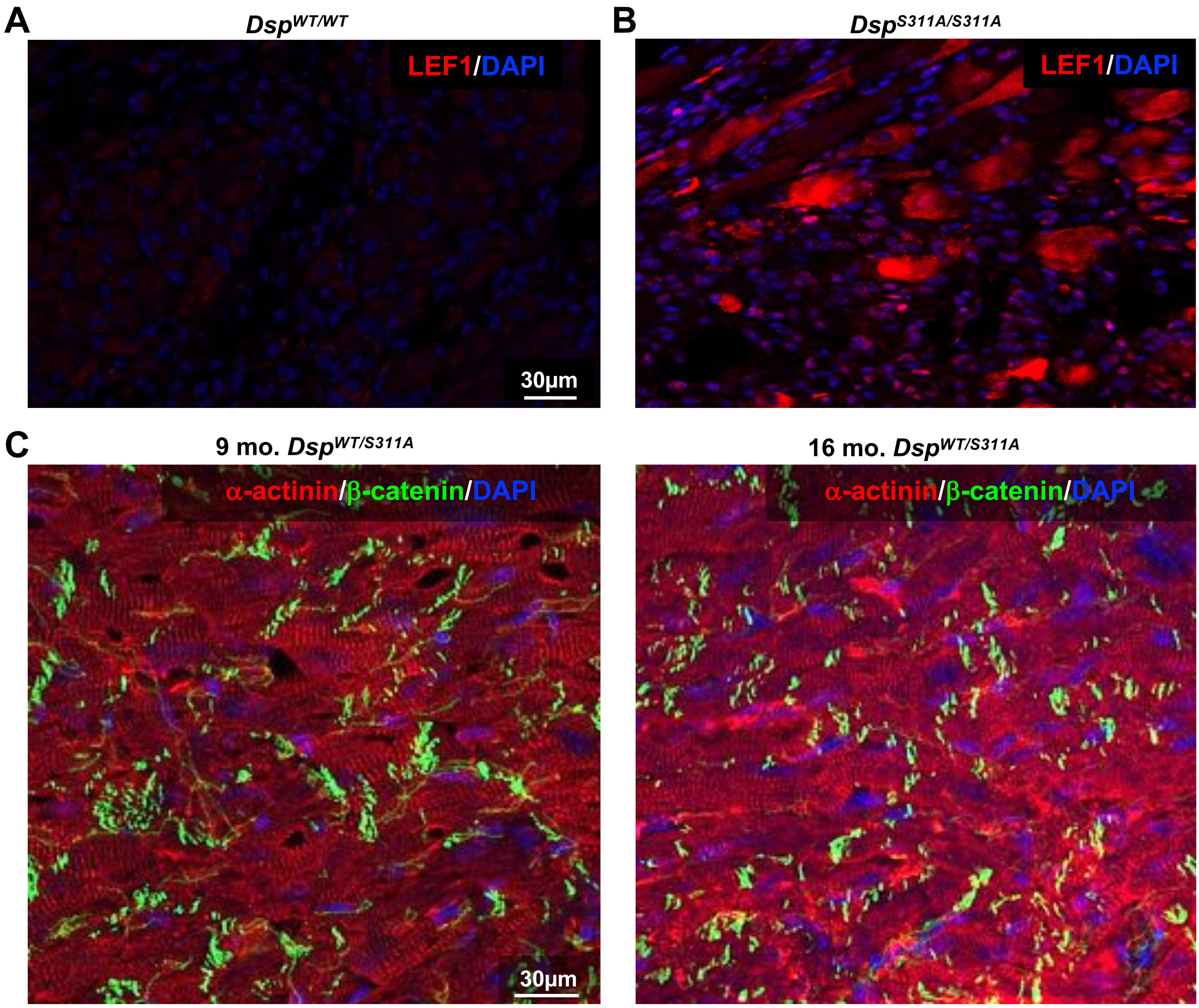

**Supplementary Figure 18**

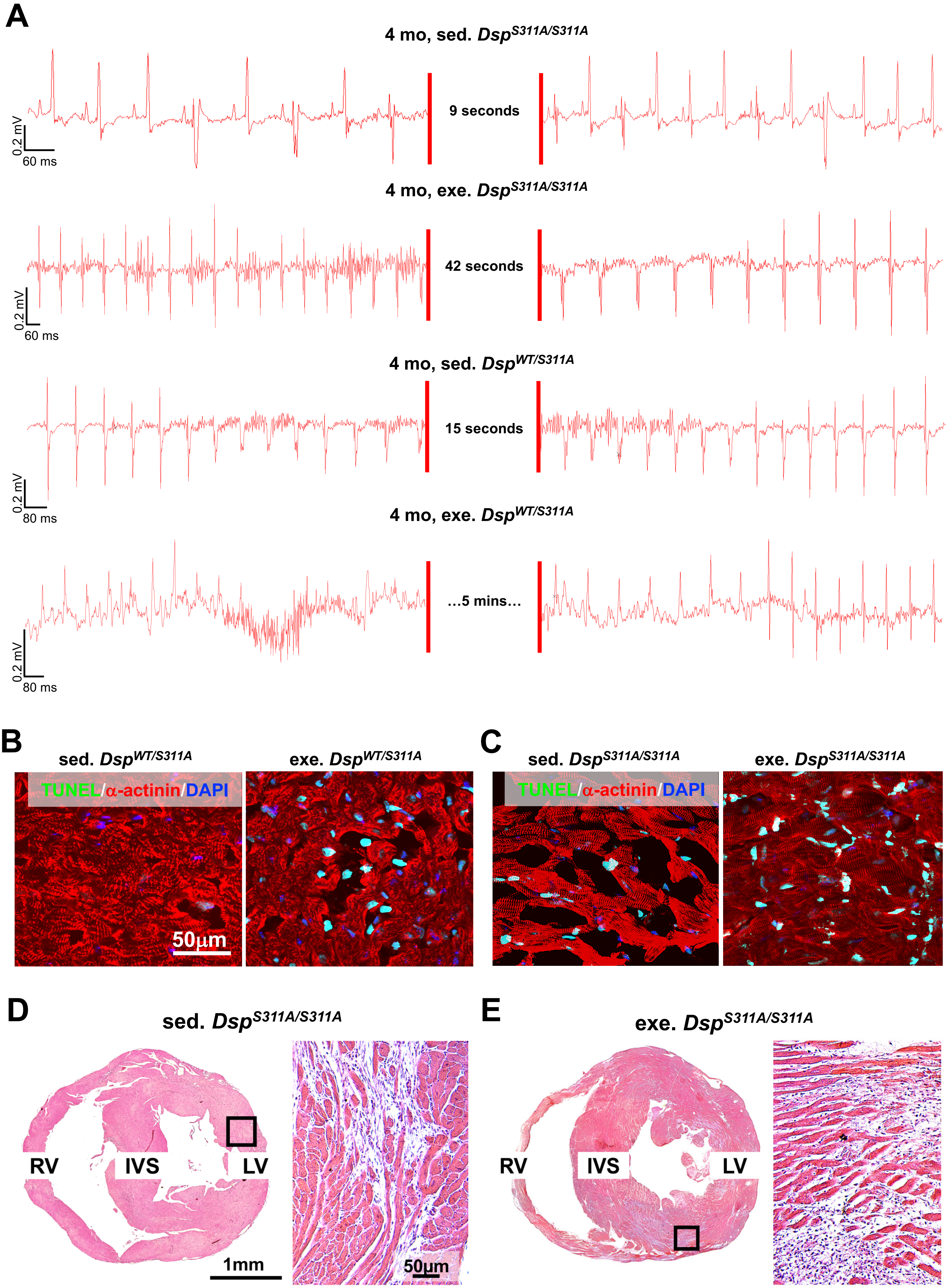

**Supplementary Figure 19**
